# AI-driven selection of patients with non-valvular atrial fibrillation for oral anticoagulation therapy: a multi-cohort validation and impact evaluation study

**DOI:** 10.64898/2026.03.23.26349067

**Authors:** Shishir Rao, Marjan Walli-Attaei, Nouman Ahmed, Zhengxian Fan, Ben O Petrazzini, Jie Lian, Seyyedhadi Ghamari, Malgorzata Wamil, Gregory Lip, Jose Leal, Kazem Rahimi

## Abstract

**Background:** Current risk assessment tools for guiding direct oral anticoagulant (DOAC) therapy for patients with atrial fibrillation (AF) based on clinical risk factors demonstrate modest predictive performance limiting clinical impact. Additionally, while guidelines recommend periodic reassessment of risk over time, there remains an absence of modelling solutions for capturing evolving risk in AF patients.

**Methods:** Using UK electronic health records, we developed and validated the Transformer-based Risk assessment survival model (TRisk), an artificial intelligence model that predicts 12-month thromboembolic and bleeding events in AF patients by leveraging temporal patient journeys up to baseline. A cohort of 411,850 prevalent non-valvular AF patients aged ≥18 years between 2010 and 2020 was identified from 1,442 English general practices. Practices were randomly allocated to derivation (n=1,079) and external validation (n=363) cohorts. TRisk was compared with CHA_2_DS_2_-VASc and CHA_2_DS_2_-VA for thromboembolic event prediction, and HAS-BLED and ORBIT for bleeding prediction, with subgroup analyses by sex, age, and baseline characteristics. A second validation of TRisk was also performed on 16,218 US AF patients between 2010 and 2023. A decision model compared outcomes and healthcare costs for TRisk versus standard care.

**Findings:** TRisk achieved higher discrimination for thromboembolic event prediction (C-index: 0.82; 95% confidence interval [CI]: [0.81, 0.83]) as compared to CHA_2_DS_2_-VASc (0.71 [0.70, 0.73]) in UK validation. Application of TRisk to US data yielded similar C-index: 0.82 (0.80, 0.84). For bleeding prediction, TRisk (C-index: 0.70 [0.69–0.71]) outperformed both HAS-BLED (0.63; [0.61, 0.64]) and ORBIT (0.64; [0.63, 0.65]), with comparable US results (0.71; [0.69, 0.74]). The model remained well-calibrated across both populations and performed equitably across subgroups, including by race and during the COVID-19 pandemic. Impact analyses showed TRisk could reduce DOAC prescriptions by 8% in the UK and 7% in the US relative to guideline-recommended approaches, while preventing at least as many thromboembolic events. This refined approach would generate annual healthcare savings of £5.5 million and $456.2 million in the UK and US respectively among patients initiating DOACs, rising to £48.6 million and $1.8 billion when extended to all AF patients on DOACs.

**Interpretation:** TRisk enabled more precise prediction for both thromboembolic and bleeding events across AF populations in UK and US compared to established clinical scoring systems. Incorporating TRisk into routine AF care would result in substantial cost savings without compromising the identification of true high-risk patients.

**Funding:** None

## Introduction

Atrial fibrillation (AF) is the most common cardiac arrhythmia, affecting over 33 million people worldwide.^1,2^ Patients with AF face up to a five-fold increased risk of thromboembolic events.^3^ With AF incidence rising globally, particularly in the UK and Europe, clinical care pressures from both AF management and subsequent thromboembolic complications are intensifying.^1,3^ To this end, preventing thromboembolic events is paramount, with direct oral anticoagulants (DOACs) serving as the cornerstone preventive therapy globally.^4–7^

In contemporary practice, initiation of DOACs in non-valvular AF patients is primarily driven by thromboembolic event risk scores. The National Institute for Health and Care Excellence (NICE) and American Heart Association (AHA) guidelines advocate use of the CHA_2_DS_2_-VASc score for guiding DOAC initiation in non-valvular AF patients,^4,5,8^ given its evidence base and validation.^9^ The European Society of Cardiology (ESC) 2024 guidelines alternatively recommend using the non-sex-adjusted CHA_2_DS_2_-VA model (Level of Evidence: C). This change reflects concerns that including sex complicates decision-making and excludes non-binary, transgender, or hormone therapy patients.^4–6^ However, as usage of DOACs can lead to bleeding events, balancing benefits against risks adds therapeutic complexity, prompting guidelines to recommend monitoring bleeding risk with complementary scores (e.g., HAS-BLED, ORBIT).^4–6,10,11^

While current risk scores have found global endorsement, there is opportunity for improved risk stratification. Multiple validation studies for these scores report discrimination metrics for status-quo models consistently below 0.8 and 0.7 for thromboembolic and bleeding events prediction respectively.^8,11–13^ Additionally, risk is not static; it evolves with age, comorbidities, and therapeutic changes.^4–6,14^ Existing models rely on cumulative factors that can only accrue over time, meaning risk scores may increase but never decrease – even when clinical circumstances improve. Consequently, guidelines recommend regular reassessment of both thromboembolism and bleeding risks.^4–6^ However, it is debatable whether these established scores are appropriate for use across the entire AF patient journey. Current clinical risk scores with minimalistic predictor sets were developed in early-stage, DOAC-naïve AF cohorts. Consequently, they capture predominantly macro-level, age-driven changes but overlook more nuanced shifts in AF patient medical profiles.^13^ Alternatively, models designed to capture evolving, multi-factorial risk would enable more accurate stratification for guiding DOAC therapy.^4–6,13,14^

The widespread availability of longitudinal electronic health record (EHR) datasets presents unprecedented opportunities for advanced risk modelling that captures evolving, multi-factorial patient profiles using routine clinical data.^15,16^ Artificial intelligence (AI) models, particularly the rise of healthcare-tailored Transformer models such as the Transformer-based Risk assessment survival model (TRisk), have shown breakthrough performance in patient health modelling.^17,18^ These AI architectures capture complete patient medical histories “from cradle to grave,” potentially identifying risk factors beyond those explicitly included in traditional scoring systems.

Motivated by the limitations of current non-valvular AF risk assessment approaches and the promise of AI-driven prognostication, we developed and validated TRisk for prediction of thromboembolic and bleeding outcomes in non-valvular AF patients across UK and US EHR datasets.

## Methods

### Overview

Using a UK cohort of AF patients, we derived TRisk for thromboembolic and bleeding event prediction and externally validated it in a separate UK cohort. In validation, we compared against benchmark models for thromboembolic (CHA_2_DS_2_-VASc, CHA_2_DS_2_-VA) and bleeding (HAS-BLED, ORBIT) events prediction.^8,10,11^ We then adapted TRisk for the US EHR setting and additional external validation was conducted. To understand TRisk’s decision-making processes, we also analysed input EHR encounters that the model identified as important for outcome prediction across cohorts. Finally, we assessed the potential impact of using TRisk to guide DOAC initiation in terms of changes to DOAC users, costs, and bleeding events.

### Data sources

For the UK data study, we used English data from the Clinical Practice Research Datalink (CPRD) Aurum dataset with approval from the CPRD Independent Scientific Advisory Committee (protocol number: 20_095).^16^ CPRD is a representative UK dataset containing detailed patient records from participating general practices, covering approximately 20% of the population. The dataset includes demographics, diagnoses, prescribed treatments, and health-related lifestyle variables, with linkage to Hospital Episode Statistics (HES) for hospital visits and Office for National Statistics (ONS) for mortality data.

For the US data study, we used EHR data from the All of Us (AoU) Research Program.^15^ AoU is a National Institute of Health initiative that aims to create a diverse biomedical dataset with over one million participants, focusing on underrepresented groups including racial minorities, rural communities, and those with lower socioeconomic status. The program gathers longitudinal EHR data from over 450,000 participants as of 2025.

### UK cohort selection and baseline definition

In UK data, we identified patients aged ≥18 years with AF during the study period. For CPRD, the study period was defined from the latest of: January 1, 2010, patient’s 18th birthday, and 12 months after GP practice registration, ending at the earliest of: last data collection, patient departure from practice, or December 31, 2019. Patients with AF before the study period were excluded.

For each patient, baseline was randomly selected from the period between first AF diagnosis and end of study period, simulating predictions at various stages of AF progression across different calendar years. This approach aims to validate the model for reliable performance not just at earlier stages of AF but for any point of dynamic assessment throughout the disease course.^18^ In this way, we investigated future risk of clinical outcomes in those with prevalent AF who have not initiated DOACs as well as residual risk in those who have initiated DOACs. Lastly, to capture a cohort that can use DOACs, patients with mitral stenosis or prosthetic valve replacement at baseline were excluded.^4–6^

### Primary and secondary outcomes

The primary outcome was thromboembolic events within 12 months, defined as ischaemic stroke, transient ischaemic attack, peripheral embolism, and pulmonary embolism identified by validated phenotyping libraries for GP, HES, or ONS records.^1,19^ Patients were censored at the earliest of the study end date or thromboembolic event date following baseline.

Separately, as a secondary outcome, we investigated major bleeding events within 12 months, defined as bleeding at critical sites (i.e., intracranial, intraspinal, intraocular, pericardial, intra-articular, intramuscular, retroperitoneal), bleeding requiring hospitalisation, or fatal bleeding identified in GP, HES, ONS records.^19^ Patients were similarly censored at the earliest of the study end date or date of bleeding event following baseline. Identification for both conditions was conducted using validated phenotyping libraries (codes in **Supplementary Data: Diagnostic codes for identifying thromboembolic and bleeding events**).^1,19^

### Validation strategy

We included data from 1,442 contributing GP practices from England, randomly assigning three-quarters (1,079 practices) to the derivation dataset and the remaining (363 practices) to external validation, following established practice-based validation methodology.^20^

### Modelling

TRisk is a Transformer survival model with regularisation procedures for well-calibrated survival modelling.^17,21^ The model processes patients’ medical histories as sequences of timestamped records, incorporating 4,687 diagnosis, 1,312 medication, 2,624 procedure, and 52 laboratory tests and measurement codes from linked primary and secondary care records totalling 8,675 codes. Relative ordering (i.e., visit information) and patient age at each encounter provide rich longitudinal annotations; EHR was utilised as-is up to baseline without imputation of missing values (**Figure S1**; modelling details in **Table S1** and **Supplementary Methods: EHR pre-processing and AI modelling**).

We compared against benchmarks, CHA_2_DS_2_-VASc and CHA_2_DS_2_-VA for thromboembolic prediction and HAS-BLED and ORBIT for bleeding events prediction.^8,10,11^ All benchmark model predictors were extracted from patient records using validated phenotyping dictionaries from previous studies.^1,12,19^ As international normalised ratio (INR) is not well recorded in CPRD, labile INR was not used for the HAS-BLED risk tool (nor would it be relevant on patients taking DOACs); instead a modified eight-point HAS-BLED score was used as in previous studies.^19^ Several variables including alcohol use, creatinine, ALT, AST, alkaline phosphatase (AP), GFR, haematocrit, haemoglobin, and bilirubin had missing values. Missing data in the validation dataset were imputed using multiple imputation by chained equations (MICE) (see **Supplementary Methods: Implementation of benchmark statistical models**).^22^

### Performance analysis

Model performance was evaluated using various metrics. Discrimination was assessed using concordance index (C-index) and area under the precision-recall curve (AUPRC), with subgroup analyses examining performance across sex, age groups, baseline comorbidities, and medication use. Calibration was evaluated graphically using smoothed calibration curves with restricted cubic splines.^23^ Benchmark models’ numerical risk scores were transformed to continuous risk scores (0-1) using Cox models, following previous methodologies.^12,19^ Decision curve analysis was performed to analyse the net benefit of the various modelling solutions.^24^

### Explainability analyses

We employed integrated gradients methodology to analyse which medical history encounters most influenced TRisk’s risk estimation of thromboembolic and bleeding events.^25^ Population-level importance was captured by averaging contribution scores across cohorts (details in **Supplementary Methods: Clarification on explainability analyses**).

### Impact analysis

Among patients not using DOACs, we conducted an impact analysis to estimate how many patients in the validation cohort – standardised to 1,000 individuals – would be recommended for DOAC prescription using TRisk compared to status-quo recommendation strategy: CHA_2_DS_2_-VASc at NICE recommended thresholds (score ≥2).^5,6^ Using these recommendation statistics, we then assessed the impact of implementing TRisk-based DOAC recommendation in the UK versus current standard care, quantifying the effect of reduced DOAC use on: (a) drug costs, (b) number of bleeding events, (c) bleeding-related healthcare costs, (d) quality-adjusted life-years (QALYs) and (e) the maximum per-patient price at which the TRisk strategy remains cost-neutral. The base case analyses were restricted to DOAC initiators; a sensitivity analysis extending to all DOAC users (both new and existing) was conducted to estimate the upper bound of the potential impact.

We also compared TRisk against CHA_2_DS_2_-VA at ESC recommended threshold (score ≥2).^6^ With these statistics, we similarly investigated impact in other European countries including Netherlands, Germany, Italy, Spain, and France. Full details of the decision model, analytic approach, assumptions, and parameter sources are provided in **Supplementary Methods: Economic impact analysis.**

### US data analysis

We conducted external validation of TRisk for 12-month thromboembolic and bleeding event prediction using the AoU US dataset as an independent cohort. Following similar cohort selection criteria used in CPRD, we included patients from January 1 2010 to January 1 2023, randomly allocating 60% to the validation dataset with the remaining 40%, forming the fine-tuning dataset (details in **Supplementary Methods: Clarification on AoU validation study**).

We used transfer learning to adapt the TRisk model to the US data context.^26^ This addressed domain shift along two axes: geographical (UK to US) and temporal (pre-2020 to 2023 data).^18,26^ The model originally trained on the CPRD derivation dataset was then tuned on the AoU fine-tuning dataset.

Since EHR patterns differ between the UK and US, we adapted TRisk by transferring existing codes and their embeddings. New AoU-specific codes were mapped to their closest ontological matches. For example, we assign the "I50" embedding to "I50.2" when unavailable (details in **Supplementary Methods: Clarification on modelling on All of Us dataset**). Following extension of our vocabulary, 9,251 codes were used to model diagnoses, medications, procedures, laboratory tests, and measurements.

As benchmark models, we used CHA_2_DS_2_-VASc and HAS-BLED for thromboembolic and bleeding event prediction respectively, in line with ACC/AHA/ACCP/HRS guidelines.^4^

We evaluated model performance on AoU validation data through discrimination, calibration, decision curve, impact, and explainability analyses. Bias analyses investigated subgroup performance focusing on sex, race, age groups, calendar year groups, baseline comorbidities, and medication use. For impact analyses, we similarly modelled status-quo clinical practice by evaluating CHA_2_DS_2_-VASc at recommended thresholds (≥2 for men, ≥3 for women) per ACC/AHA/ACCP/HRS guidelines for AF thromboembolic event prevention.^4^

In supplementary analysis, we compared two additional TRisk variants against the transfer learning approach: one trained solely on CPRD data (conventional external validation) and another trained solely on the AoU fine-tuning dataset (internal validation). Detailed modelling information is provided in **Supplementary Methods: Clarification on modelling on AoU dataset**.

### Reporting guidelines

We followed TRIPOD-AI reporting guidelines for transparent reporting of our study.^27^

## Results

### UK cohort analyses

From UK CPRD data (**Figure S2**), 411,850 patients (47% female) were selected for analysis (98,176 in validation) with median follow-up of 8 (interquartile interval [IQI]: 2-30) months. Median age was 79 (IQI: 70-86) years. Approximately a quarter of patients had diabetes or previous stroke events, and 71% were free of DOAC use at baseline (**Table 1**). Thromboembolic events occurred in 6.9% of patients within 12 months, while major bleeding occurred in 7.1%.

**Table 1.** UK population characteristics.

| Characteristic | Derivation | Validation |
| --- | --- | --- |
| Patients (%) | 313,674 (76.2%) | 98,176 (23.8%) |
| Women (%) | 148,973 (47.5%) | 46,272 (47.1%) |
| Median age (years) (IQI) | 79.0 (70.0, 86.0) | 79.0 (70.0, 86.0) |
| Median time since AF diagnosis (months) (IQI) | 8.4 (1.7, 24.1) | 8.4 (1.7, 24.1) |
| Median SBP (mmHg) (IQI)† | 133.9 (125.4, 142.2) | 134.0 (125.5, 142.4) |
| Median creatinine (μmol/L) (IQI)† | 95.0 (79.0, 118.0) | 95.0 (79.0, 118.0) |
| Median bilirubin (μmol/L) (IQI)† | 12.0 (9.0, 16.0) | 11.0 (9.0, 16.0) |
| Drinking status† |  |  |
| Non-drinker (%) | 158,546 (50.5%) | 49,839 (50.8%) |
| Very light drinker (<1 units/day) (%) | 85,389 (27.2%) | 26,773 (27.3%) |
| Light drinker (1-2 units/day) (%) | 46,838 (14.9%) | 14,447 (14.7%) |
| Moderate drinker (3-6 units/day) (%) | 15,342 (4.9%) | 4,824 (4.9%) |
| Heavy drinker (7-9 units/day) (%) | 2,985 (1.0%) | 908 (0.9%) |
| Very heavy drinker (>9 units/day) (%) | 4,574 (1.5%) | 1,385 (1.4%) |
| Disease history |  |  |
| Stroke/thromboembolism (%) | 77,607 (24.7%) | 24,437 (24.9%) |
| MI/PAD/aortic plaque (%) | 83,098 (26.5%) | 26,214 (26.7%) |
| Diabetes (%) | 64,547 (20.6%) | 21,035 (21.4%) |
| Hypertension (%) | 250,237 (79.8%) | 78,636 (80.1%) |
| Congestive heart failure (%) | 47,516 (15.1%) | 15,006 (15.3%) |
| Liver disease (%) | 16,909 (5.4%) | 5,229 (5.3%) |
| CKD (%) | 97,807 (31.2%) | 30,591 (31.2%) |
| Previous bleeding (%) | 115,278 (36.8%) | 36,155 (36.8%) |
| Prescription history |  |  |
| Direct oral anticoagulants (%) | 89,603 (28.6%) | 28,282 (28.8%) |
| VKAs (%) | 71,274 (22.7%) | 23,012 (23.4%) |
| Aspirin, clopidogrel, NSAIDs (%) | 273,782 (87.3%) | 87,536 (89.2%) |
| *SBP: systolic blood pressure; CKD: chronic kidney disease; PAD: peripheral arterial disease; MI: myocardial infarction; NSAIDs: non-steroidal anti-inflammatory drugs; VKAs: Vitamin K antagonists.*<br>*<sup>†</sup>indicates missingness of following variables: SBP: 27,184 (6.6%) patients; creatinine: 49,763 (12.1%); bilirubin: 49,857 (12.1%); drinking status: 350,452 (85.1%).* |  |  |

**Table 2.**
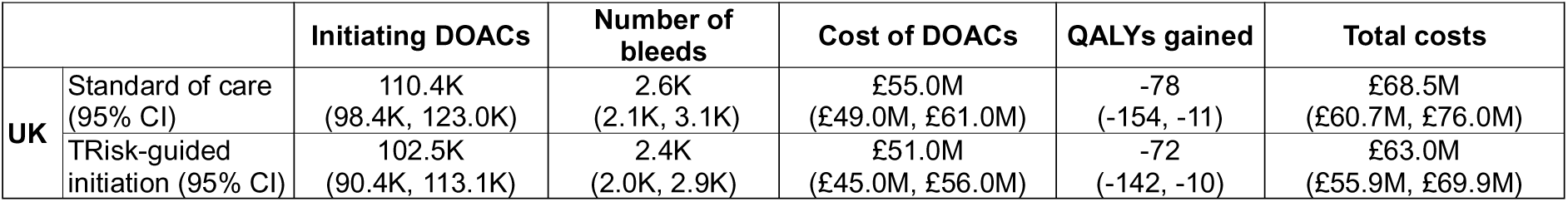
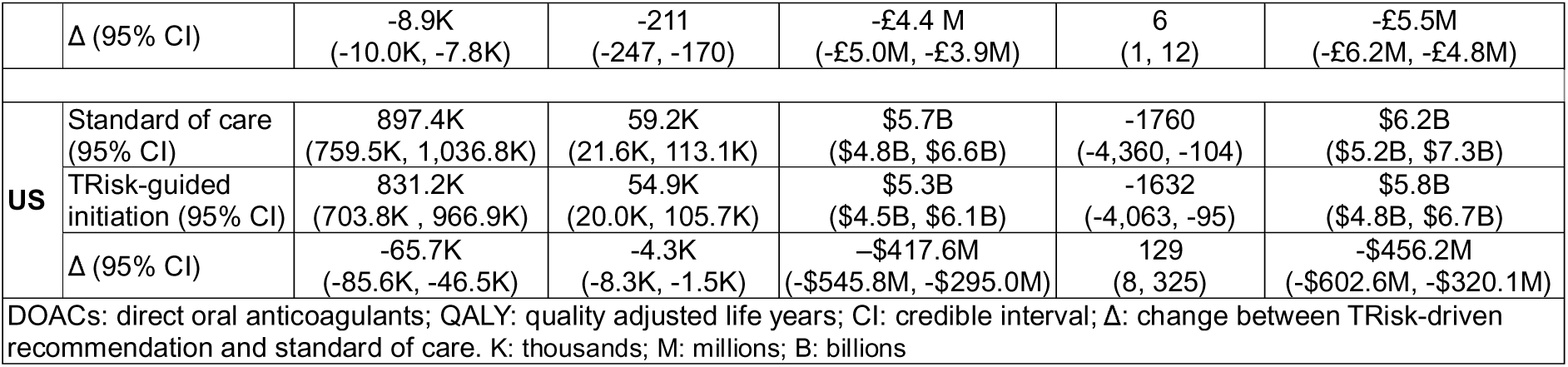
Impact of TRisk-guided recommendation of direct oral anticoagulants as compared to standard of care in UK and US.

TRisk achieved improved discrimination for thromboembolic event prediction with C-index 0.818 (95% CI: 0.808 - 0.828) compared to CHA_2_DS_2_-VASc (0.713; [0.701 - 0.725]) and CHA_2_DS_2_-VA (0.717; [0.705 - 0.728]) – a 13% improvement on both benchmark risk scores (**Figure 1**). AUPRC findings were similar with TRisk achieving 0.229 versus 0.123 for CHA_2_DS_2_-VASc and CHA_2_DS_2_-VA (**Table S2**).

**Figure 1.**
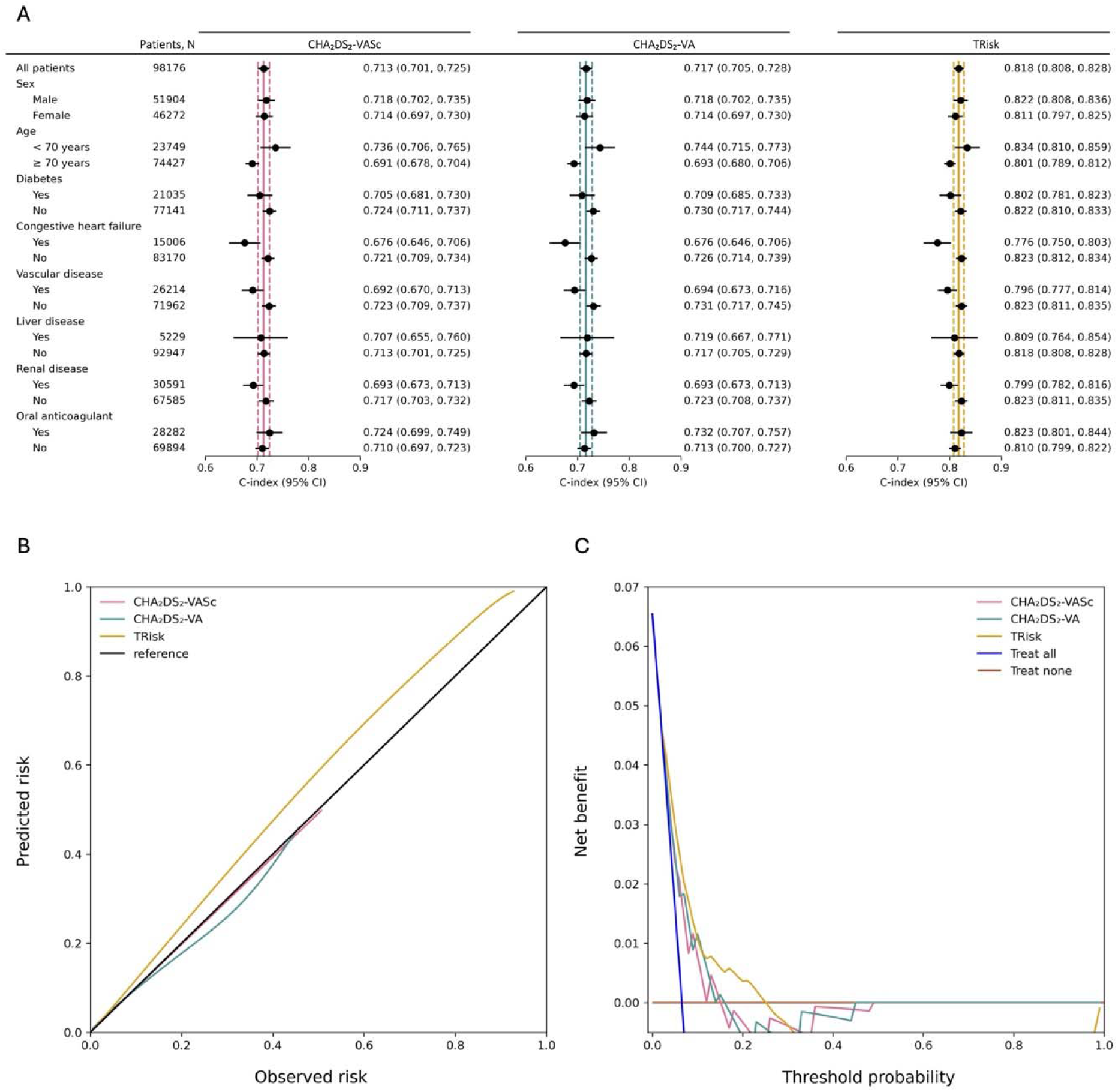
Discrimination, calibration, and clinical utility of thromboembolic risk models in the UK validation cohort. (A) Models’ discrimination by concordance index (C-index) with 95% confidence intervals (CI) in the overall cohort and subgroups for 12-month thromboembolic event risk prediction. Maroon, teal, and gold lines represent the C-index for all patients in the validation cohort for CHA_2_DS_2_-VASc, CHA_2_DS_2_-VA, and TRisk, respectively. Solid and dotted lines show the mean and 95% CI of the overall cohort C-index for each model and are provided to visually demonstrate deviation in subgroup performance from overall cohort performance. SBP: systolic blood pressure. (B) Calibration curves for all models. The reference line (black) indicates perfect calibration. (C) Net benefit curves for all models. The x-axis shows the decision threshold probability, and the y-axis shows net benefit, defined as the difference between the proportion of true positives and false positives, weighted by the odds of the chosen decision threshold.

Subgroup analyses showed that TRisk consistently outperformed benchmark scores across all strata, with no material deviation from overall discrimination. C-index remained >0.8 even in patients not using DOACs at baseline (**Figure 1A**).

TRisk and comparators were acceptably calibrated across clinically relevant risk thresholds. Decision curve analysis demonstrated substantially greater net benefit for TRisk between thresholds 0.0–0.3 (**Figure 1B,1C**).

For bleeding events prediction, TRisk achieved C-index 0.701 (0.690 - 0.712), outperforming HAS-BLED (0.625; [0.613 - 0.636]) and ORBIT (0.638; [0.626 - 0.650]) scores (**Figure S3**). TRisk similarly outperformed benchmarks in terms of AUPRC (**Table S2**); similar performance was maintained in patients not using DOAC across models (**Figure S3**). Finally, TRisk demonstrated good calibration in the risk spectrum of interest and provided greater net benefit than benchmark models (**Figure S4**).

### US cohort analyses

The AoU dataset included 16,218 non-valvular AF patients (9,731 in validation) with median follow-up of 14 months for thromboembolic and bleeding events. There were 2,096 (12.9%) thromboembolic and 2,196 (13.5%) bleeding events in the first year (**Figure S5; Table S3**).

The transfer-learning TRisk variant achieved a C-index of 0.822 (0.800–0.844) for thromboembolic prediction, outperforming CHA_2_DS_2_-VASc (0.722 [0.696–0.749]) with appropriate calibration and greater net benefit and AUPRC (**Figures 2, S6–S7; Table S4**). Importantly, there was no evidence of bias across subgroups - including sex, race, age, and the COVID-19 pandemic era (baseline year after 2020).

**Figure 2.**
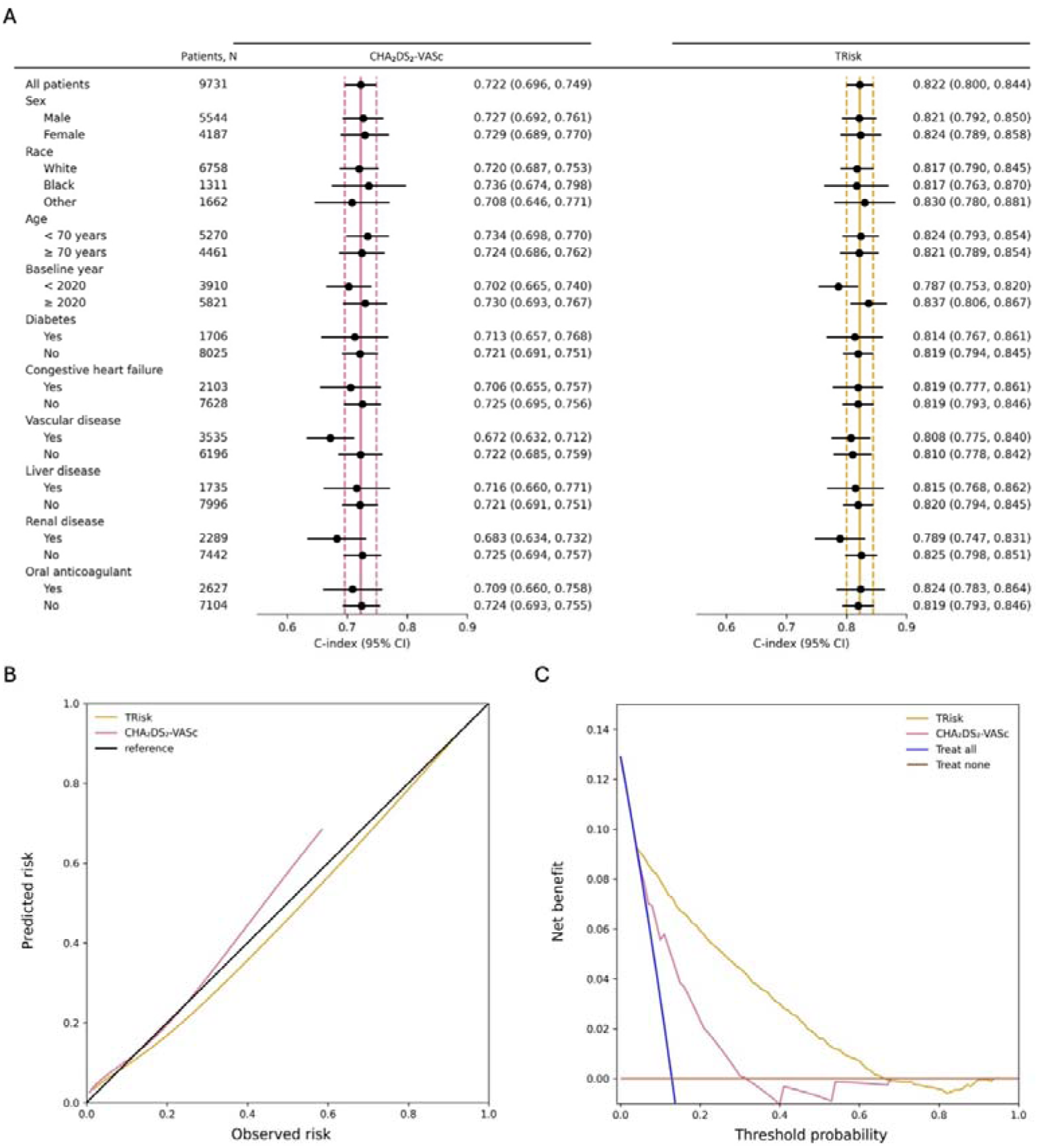
Discrimination, calibration, and clinical utility of thromboembolic risk models in the US validation cohort. (A) Models’ discrimination by concordance index (C-index) with 95% confidence intervals (CI) in the overall cohort and subgroups for 12-month thromboembolic event risk prediction. Maroon, teal, and gold lines represent the C-index for all patients in the validation cohort for CHA_2_DS_2_-VASc, CHA_2_DS_2_-VA, and TRisk, respectively. Solid and dotted lines show the mean and 95% CI of the overall cohort C-index for each model and are provided to visually demonstrate deviation in subgroup performance from overall cohort performance. SBP: systolic blood pressure. (B) Calibration curves for all models. The reference line (black) indicates perfect calibration. (C) Net benefit curves for all models. The x-axis shows the decision threshold probability and the y-axis shows net benefit, defined as the difference between the proportion of true positives and false positives, weighted by the odds of the chosen decision threshold.

TRisk also surpassed models trained solely on UK data or *de novo* on AoU and was well-calibrated (**Figure S7**). For bleeding event prognostication, TRisk achieved a C-index of 0.714 (0.690–0.738) versus 0.658 (0.632–0.683) for HAS-BLED. Similarly, TRisk remained well-calibrated, provided greater net clinical benefit, and showed no evidence of bias across sex, race, age groups, or other variables (**Figures S8–S9; Table S4**).

### Explainability analyses

Integrated gradients analysis on TRisk modelling identified consistently important factors across CPRD and AoU cohorts (**Figure 3**). Previous thromboembolic, stroke, and cardiovascular events contributed to increase in risk scores in both UK and US analyses. Conversely, established DOACs (i.e., rivaroxaban, apixaban) contributed to attenuation in thromboembolic risk scores. In patients not taking DOACs, similar patterns were found for factors contributing to elevated thromboembolic event risk score (**Figure S10**).

**Figure 3.**
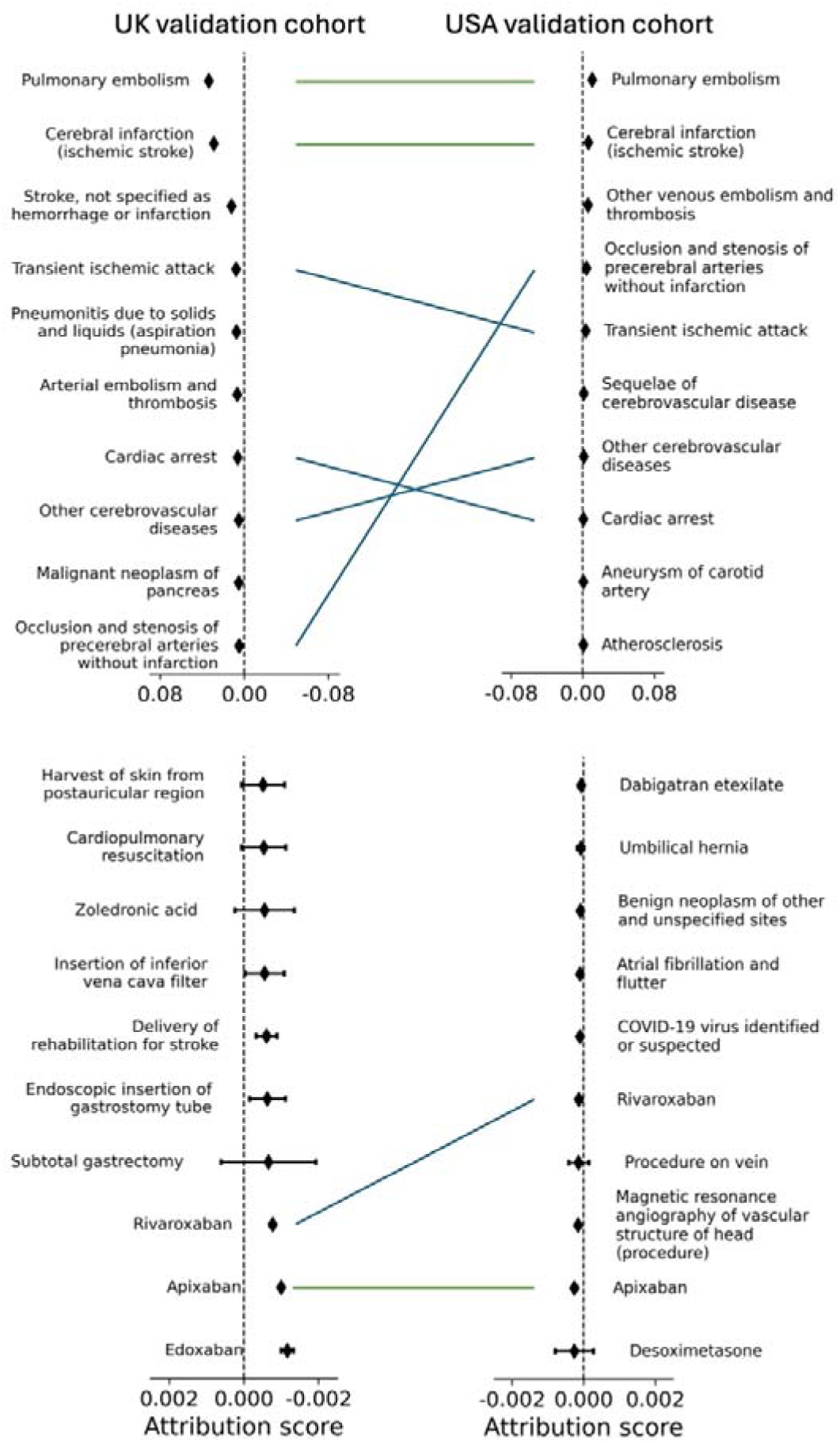
Average contribution for most positively (above) and negatively (below) contributing ten encounters for thromboembolic event risk prediction on UK and US validation cohort datasets. Contribution score measures how much each encounter contributes to the model’s final prediction. Contribution scores are captured by integrated gradient method for the TRisk model for prediction of thromboembolic events across UK (left) and US (right) datasets. Green line denotes both encounter and rank were preserved between validation analyses; blue line denotes only encounter was preserved. Please note different scales for top and bottom sets of forest plots.

In analyses of bleeding event prediction, only two medical history events showed exact overlap as risk contributors between cohorts: intracranial injury and diseases of the digestive system (**Figure S11**). However, broader thematic overlap existed among bleeding-related risk contributors (e.g., various haemorrhages and diseases of the digestive system). Similar patterns were observed in patients not taking DOACs (**Figure S12)**.

### Impact analyses

For clinical utility, we reasoned that any new model must capture at least as many patients who will experience thromboembolic events as standard care. We therefore selected the TRisk threshold to match CHA_2_DS_2_-VASc true positive rates. At a 3% threshold, TRisk identified the same number of thromboembolic events as CHA_2_DS_2_-VASc at the NICE-recommended threshold (71 true positives; 2 false negatives per 1,000 patients) while recommending DOACs for 743 rather than 814 patients (8% reduction) (**Figure 4A; Table S5**).^5^ Compared with CHA_2_DS_2_-VA at the ESC-recommended threshold, TRisk still recommended 41 fewer patients (5% reduction) and captured one additional true positive (**Table S5**).^6^

**Figure 4.**
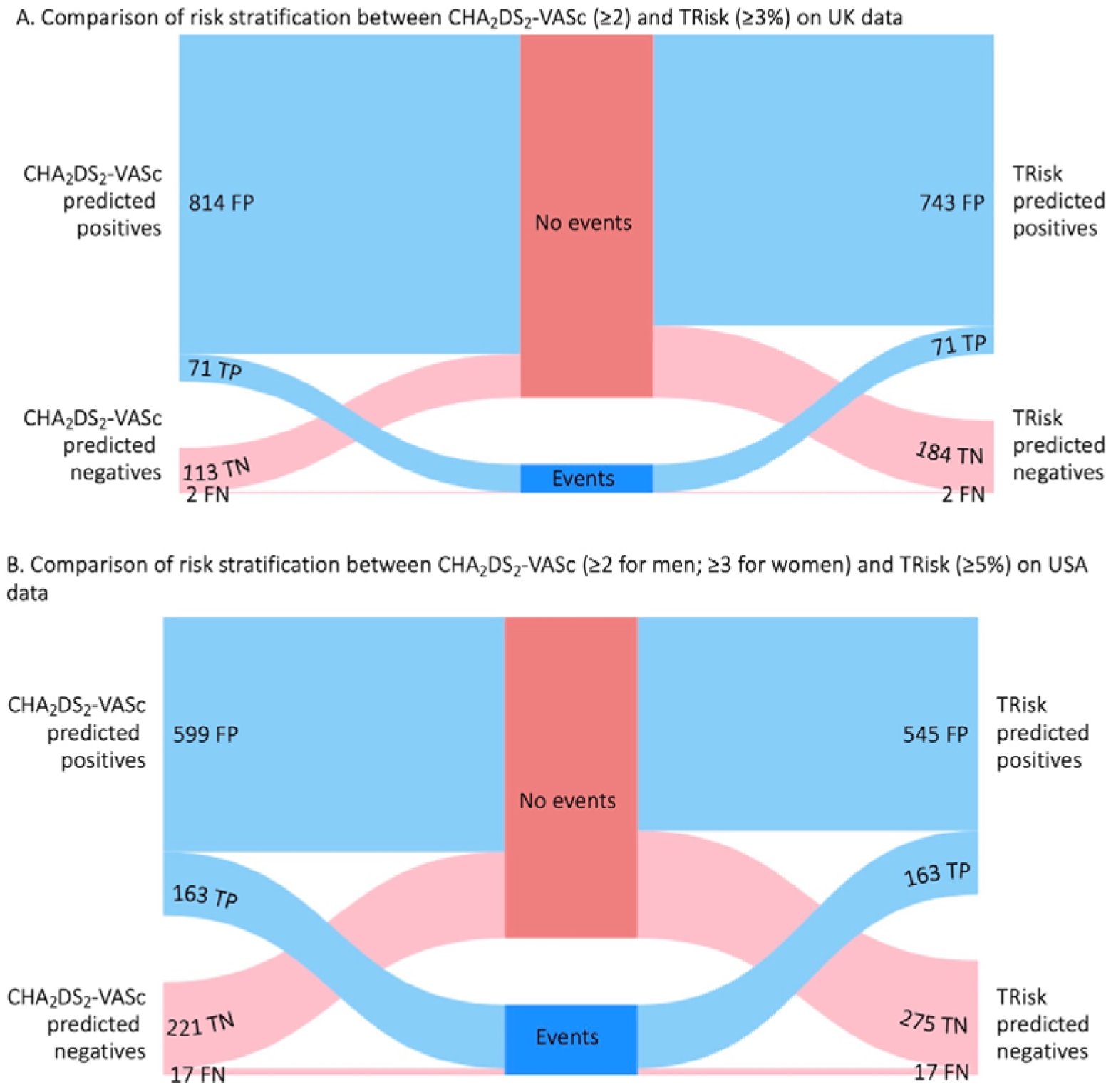
Impact of various strategies for guiding direct oral anticoagulant (DOAC) recommendations in atrial fibrillation patients free of DOAC use on UK and US data. (A) The figure illustrates true positives (TP), false positives (FP), true negatives (TN), and false negatives (FN) capture for CHA_2_DS_2_-VASc (left) versus TRisk (right) on (A) UK and (B) US data. In UK data, CHA_2_DS_2_-VASc is presented at threshold ≥2, TRisk at threshold ≥3%. In US data, CHA_2_DS_2_-VASc is presented at threshold ≥2 for men; ≥3 for women, TRisk at threshold ≥5%. To aid presentation, all numbers presented are standardised to cohort of 1,000 patients who have not initiated oral anticoagulants.

Scaled to the UK untreated AF population (**Tables 2, S6-S9; Figure S13**), TRisk would avoid DOAC initiation in 8,857 patients (95% CI: [7,822-10,005]) annually, preventing 211 bleeding events (170-247), saving £5.5 million in total healthcare costs (£4.8-£6.2 million) and gaining 6 QALYs (1-12). Cost-neutrality is preserved at implementation costs up to £50 per patient (£45-£54). Extending to all DOAC users, projected savings increase to £48.6 million (£42.5–£54.6 million), preventing 1,867 bleeding events and gaining 55 QALYs (3-106). Across five exemplar European countries, a 5% reduction in selection of at-risk patients for DOACs via TRisk among 5.2 million patients with AF could save approximately €17.0 million (€15.0-€19.0) per year among DOAC initiators, rising to €178.3 million (€162.0-€193.7 million) when extended to all DOAC users (**Tables S8-S10**).

In US data, TRisk at a 5% threshold recommended 54 fewer patients per 1,000 (7.2% reduction) than CHA_2_DS_2_-VASc at AHA thresholds,^4^ with identical true positives (**Figure 4B; Table S11**). Scaled to the US untreated AF population, TRisk would safely reduce DOAC initiation by 65,688 patients annually (46,481–85,594) in the base case, preventing 4,335 bleeding events (1,537-8,337) and saving $456.2 million ($320.1–$602.6 million). Cost-neutrality is preserved at implementation costs up to $508 per patient ($362-$650). When extended to all DOAC users, TRisk would deprescribe for 263,323 patients (198,880-329,456), preventing 17,379 bleeding events (6,303–35,469) and saving $1.8 billion ($1.4-$2.3 billion) annually (**Tables S8-S11**).

## Discussion

Using routine EHR data, TRisk achieved C-indices exceeding 0.8 and 0.7 for thromboembolic and bleeding events respectively while maintaining appropriate calibration across two large-scale international datasets. The model demonstrated robust prognostication without bias across subgroups – particularly by sex, race and in different time periods (e.g., COVID-19 pandemic). Explainability analyses captured risk factors both established and less appreciated. TRisk would safely reduce DOAC recommendations by 8% in the UK and 7% in the US compared to guideline-endorsed strategies, while capturing just as many thromboembolic events. This would translate into total potential cost savings of £5.5 million and $456.2 million per year among DOAC initiators, rising to £48.6 million and $1.8 billion when extended to all DOAC users in the UK and US respectively.

Rising AF incidence globally puts pressure not only on AF management but also on managing its sequelae, particularly thromboembolic complications.^1,2^ Current thromboembolism risk scores show modest predictive performance: CHA_2_DS_2_ scores achieve discrimination below 0.8, and bleeding scores below 0.7. Our large-scale EHR implementation confirmed these limitations, consistent with previous validation studies.^12,19^

Furthermore, while clinical guidelines strongly recommend routine reassessment, few solutions exist for robust dynamic risk evaluation throughout the AF patient journey. Indeed, while clinical risk scores change over time, the nuance is limited; studies show most risk progression results from aging past 65 and 75 years (adding one point each).^13,28^ Hence, it is unsurprising that reassessment protocols are not followed consistently, with studies showing that only one-fifth of GPs conduct OAC prescription reassessment via established clinical risk scores.^5,6,29^ Alternatively, models explicitly designed to capture evolving multi-factorial risk independent of age would better align with guideline frameworks and enable more precise risk stratification for optimising DOAC therapeutic decisions.^4–6,13^

To achieve this, we explicitly designed the study to test models’ ability to perform risk assessment throughout the AF patient journey. By randomly selecting baseline timepoints within eligible assessment periods, we ensured the cohort represented patients at all disease stages rather than skewing toward early or late AF phases.^30^ Within this framework, we developed and validated TRisk against established benchmarks for both thromboembolic and bleeding event prediction.

TRisk’s superior performance on both prognostication tasks stems from capturing complex patient journeys through comprehensive longitudinal health data analysis. Unlike traditional snapshot assessments, TRisk processes entire medical histories – diagnoses, medications, procedures, and laboratory results – automatically identifying patterns that conventional approaches miss. This data-driven methodology proves particularly valuable for conditions like AF where traditional risk factors explain only partial outcome variance. With this generalisable framework, TRisk materially outperformed benchmarks for both thromboembolic and bleeding event prediction.

Explainability analyses highlighted TRisk’s ability to capture complex patient profiles. It recovered established risk and protective factors^2^ while identifying novel predictors absent from traditional scores, such as cardiac arrest. These factors were consistently important across both validation cohorts, suggesting genuine risk effects rather than dataset artefacts. TRisk also reflected evolving risk profiles: apixaban and rivaroxaban attenuated risk in the overall cohort, while other encounters did so in patients not on DOACs. This shows risk contributions vary throughout the patient journey. Models like TRisk, capable of capturing such changes, are better positioned to evaluate dynamic risk.

Indeed, the successful transfer learning implementation from UK to US healthcare setting revealed TRisk’s potential for broader clinical applicability. Importantly, in the US cohort, we showed not just regional generalisability but also temporal generalisability. TRisk performed equally well from 2010-2020 as when validated beyond 2020 through 2023, showing that models trained on complex patient cohorts like those with AF can be successfully transferred to modern temporal periods for continued trusted use. This was achieved through a modular expansion approach: US-specific codes were mapped to their closest UK equivalents and their embeddings transferred, allowing TRisk to operate in the US setting with minimal retraining on fewer than 6,500 patients. To our knowledge, TRisk is the first risk model with this property; its AI formulation allows adaptation to new settings simply by plugging in additional codes, scalable from zero codes to thousands, making it highly practical across regions and evolving coding systems.

The impact analyses underscore the potential system-level value of TRisk. Our model maintains the same true positive capture as CHA_2_DS_2_-VASc while substantially reducing false positives. In the UK, this would safely deselect about 1 in 12 treated patients. Scaled to the UK population, this would translate to annual savings of ∼£5.5 million in DOAC and bleeding-related NHS costs^31,32^ among DOAC initiators, rising to ∼£48.6 million when extended to all DOAC users. In the US, where DOAC uptake and prices are higher, TRisk would deselect ∼1 in 14 treated patients, yielding ∼$456.2 million in annual savings among DOAC initiators, rising to ∼$1.8 billion when extended to all DOAC users. This larger absolute figure reflects both higher per-patient DOAC costs (around three-to-five times European tariffs) and greater treatment penetration.^32,33^ Collectively, these findings indicate that TRisk can deliver substantial cost savings across diverse healthcare systems while preserving and even improving clinical outcomes through fewer bleeding events – all directly aligning with global directives for cost containment and evidence-based precision therapy.

The economic case for implementation is compelling: cost-neutrality is preserved at implementation costs up to £50 per patient in the UK and $508 per patient in the US. Crucially, since TRisk simultaneously reduces costs and gains QALYs, it strictly dominates standard care under both NICE’s £25K-£35K per QALY willingness-to-pay threshold and analogous US benchmarks (∼$100K-$150K per QALY gained). Unlike most health interventions, which improve outcomes only at additional cost, TRisk is comparatively rare: it achieves better outcomes while also reducing costs.

### Strengths and limitations

This study’s strengths include comprehensive validation across multiple healthcare settings (geographical and temporal), rigorous comparison against established benchmarks, successful transfer learning, detailed explainability analyses, and comprehensive impact analyses. Large sample sizes and focus on under-represented minorities in the US data validation provide robust evidence for TRisk’s performance advantages. Regarding limitations, HAS-BLED score was not implemented as designed in UK data due to missing INR measurements in CPRD – an omission unlikely to affect DOAC-treated patients, in whom INR monitoring is unnecessary. However, all variables were available and used in AoU validation.

TRisk requires complete EHR access, which increases implementation complexity relative to traditional models. However, robust tools exist for AI deployment in resource-limited settings, and future efforts should explore TRisk as an offline tool for periodic risk estimation to avoid adding point-of-care burden.^34^ Implementation could leverage existing infrastructure; for example, the NHS’s planned "Single Patient Record" initiative, which aims to consolidate primary and secondary care data as part of its 10-year plan, could provide the necessary data infrastructure to facilitate TRisk deployment.^35^ Implementation of TRisk would simultaneously improve audit of thromboembolic event prevention services through population-level analysis while delivering individualised patient insights. Most importantly, TRisk would be seamlessly integrated with routine EHR without overwhelming healthcare professionals at the point-of-care.^1,2,34^

## Conclusion

TRisk delivers accurate, unbiased, and dynamic risk assessment for both thromboembolic and bleeding events in AF patients, validated across UK and US populations. Applied to DOAC prescribing, TRisk safely selects 8% fewer patients in the UK and 7% in the US than guideline-endorsed strategies without missing a single additional thromboembolic event. This dual dividend: fewer DOAC recommendations and bleeding complications, translates to £5.5 million and $456.2 million in annual savings among DOAC initiators alone. This rises to £48.6 million and $1.8 billion when extended to all DOAC users in UK and US respectively – transforming AF management while enhancing clinical safety.

## Data Availability

The codebase for Transformer-based Risk Assessment survival modelling is available online: https://process.innovation.ox.ac.uk/software.

## Supplementary Material

### Supplementary Methods

#### Supplementary Methods: Clarification on CPRD validation study

This research utilized patient data from multiple general practices to create two distinct datasets: one for model development (derivation) and another for testing (validation). The derivation dataset served to build and train models, while the validation dataset provided an independent test of model performance.

For the TRisk model, special precautions were implemented to prevent overfitting–a common problem in complex AI systems where models become too specialized to training data. Five percent of the derivation data was randomly set aside for end-of-epoch evaluation. This subset underwent testing after each complete training cycle, when the AI system had processed all designated training patients. This approach helped monitor whether the model was genuinely learning patterns or simply memorizing specific data points.

Once the AI model demonstrated convergence on the remaining 95% of derivation patients, its performance was assessed using the separate validation dataset. This two-stage validation process – internal monitoring during training followed by external validation – ensured robust model evaluation.

In contrast, conventional statistical models utilized the complete derivation dataset for parameter estimation, reflecting the different computational approaches and overfitting risks between traditional statistical methods and modern AI techniques. This methodology provided a comprehensive framework for comparing model performance across different analytical approaches.

#### EHR pre-processing and AI modelling

Our AI modeling approach required comprehensive standardization of all medical data modalities, including diagnoses, medications, procedures, and laboratory measurements. This standardization process converted diverse medical information into uniform "codes" that could effectively process both UK and US healthcare data, ensuring cross-system compatibility and consistent model performance.

Diagnostic codes underwent systematic transformation from GP-recorded CPRD “Medcode” formats to standardized level 4 ICD-10 classifications. This conversion maintained precision by preserving one decimal place (such as N29.9, J12.2, I50.1). As the hospital records were already in level 4 ICD format, this process created uniformity between hospital and general practice records. The transformation leveraged established phenotyping dictionaries from NHS Digital and SNOMED-CT systems, ensuring clinical accuracy and international compatibility.

Medication data processing involved mapping native CPRD "product codes" into the Observational Medical Outcomes Partnership (OMOP) framework using RxNorm vocabulary’s "ingredient" classification system. This approach focused on active pharmaceutical ingredients rather than brand names – for instance, "Jantoven" was converted to its active ingredient "warfarin." Multi-ingredient medications were systematically decomposed into constituent components, with "Acetaminophen/Codeine Oral Tablet" separated into "codeine" and "acetaminophen." Dosage information was intentionally excluded due to significant data missingness across all study datasets.

Procedural data transformation mapped Office of Population Censuses and Surveys (OPCS) Classification version 4 codes into SNOMED terminology. To ensure statistical validity and computational efficiency, only codes appearing in at least 0.1% of the CPRD Aurum database were included in the modelling process.

Understanding TRisk’s development requires examining its predecessor, BEHRT, which comprises three fundamental components. The embedding layer incorporates three distinct data streams from electronic health records. A Transformer-based feature extractor processes temporal healthcare data using sophisticated multi-head self-attention mechanisms to identify complex patterns and relationships. Finally, a binary prediction layer employs sigmoid activation functions followed by linear transformations of patient representation vectors to generate individual risk assessments.

The embedding system encompasses three critical elements: encounter data, age information, and positional encoding. Encounter embeddings capture comprehensive medical histories including diagnoses, medications, and procedures up to the baseline measurement point. Age embeddings specify exact patient age during each medical encounter, calculated by subtracting event dates from estimated birth dates. Due to pseudonymization requirements, CPRD provides only birth years, necessitating July 1st as the standardized birth date assumption. Positional encoding provides sequential context through pre-established embeddings that indicate visit numbers – first visits receive position encoding for visit 1, subsequent visits for visit 2, and so forth. These three embedding types combine to create comprehensive high-dimensional vector representations of individual patient encounters.

TRisk introduces several critical modifications to the underlying BEHRT architecture, transforming it into a sophisticated survival prediction model. The enhanced age embedding system now explicitly incorporates baseline age to capture temporal prediction contexts more effectively. This modification ensures that risk predictions for patients at age 61 differ meaningfully from those at age 65, with baseline age serving as an explicit predictive variable.

The embedding integration process represents a fundamental architectural change. Rather than summing embedding layers as in previous BEHRT iterations, TRisk concatenates the three separate embedding layers using tensor operations and applies weight vector transformations that convert the dimensional space from 3•E to E (where E represents individual embedding layer space). This concatenated result undergoes non-linear hyperbolic tangent functional transformation, enabling more expressive latent representations of raw electronic health record data while simultaneously mitigating overfitting risks.

TRisk’s most significant innovation lies in adopting the Scalable Continuous-Time Survival Model through Ordinary Differential Equation Networks (SODEN) framework. Traditional maximum likelihood estimation approaches require computationally expensive integral calculations for training on censored survival data. SODEN reframes maximum likelihood estimation as differential-equation constrained optimization, dramatically improving computational efficiency.

Unlike conventional proportional hazard frameworks that lack theoretical robustness for stochastic gradient descent using mini-batching techniques, SODEN addresses scalability challenges inherent in "big data" applications. Previous survival modeling approaches struggle with random non-overlapping subset processing from derivation datasets. SODEN’s ordinary differential equation approach to time-to-event distribution modeling creates more flexible maximum likelihood estimation frameworks for censored data, eliminating restrictive structural assumptions about survival and hazard distribution shapes while enabling scalable stochastic gradient descent training.

The final TRisk innovation involves implementing an Explicit Calibration method within the objective function. This approach optimizes D-calibration – a comprehensive calibration measure – alongside empirical loss minimization. This dual optimization strategy ensures that TRisk maintains both discrimination accuracy and calibration precision, critical factors for clinical decision-making applications.

Similar to BEHRT, TRisk utilizes a specialized "Predict token" that functions as a dense patient representation vector. This comprehensive vector captures individual patient characteristics and medical histories in a compact, computationally efficient format. The dense feature vector feeds directly into the SODEN survival modeling network, which conducts outcome-specific risk predictions for individual patients.

These architectural and methodological enhancements position TRisk as theoretically superior for large-scale electronic health record analysis. The model demonstrates enhanced expressiveness and flexibility compared to traditional approaches, ensuring optimal discrimination between high-risk and low-risk patients while maintaining accurate calibration across diverse patient populations. This combination of scalability, accuracy, and calibration makes TRisk particularly suitable for real-world clinical implementation where both predictive performance and reliability are essential for effective patient care and resource allocation decisions.

#### Implementation of benchmark statistical models

Our study implemented several established clinical prediction models as benchmarks. CHA_2_DS_2_-VASc incorporates congestive heart failure, hypertension, age categories (≥75 years, 65-74 years), diabetes, previous stroke/TIA, vascular disease, and sex. CHA_2_DS_2_-VA uses identical predictors excluding sex. HAS-BLED assesses hypertension, renal disease (dialysis, transplant, or creatinine >2.26 mg/dL/200 μmol/L), liver disease (cirrhosis or bilirubin >2x normal with AST/ALT/AP >3x normal), prior stroke, bleeding history, age ≥65, bleeding-predisposing medications (aspirin, clopidogrel, NSAIDs), and alcohol use. ORBIT evaluates haemoglobin levels (<13 g/dL males, <12 g/dL females) or haematocrit (<40% males, <36% females), age >74 years, bleeding history, reduced estimated GFR (<60 mL/min/1.73m²), and antiplatelet treatment.

All benchmark predictors were extracted using established phenotyping dictionaries. INR was excluded due to poor CPRD recording quality. Multiple variables including alcohol use, creatinine, liver enzymes (ALT, AST, AP), GFR, haematocrit, haemoglobin, and bilirubin experienced significant missingness, necessitating multiple imputation by chained equations (MICE) for CPRD validation datasets.

The imputation process included all model variables alongside Nelson-Aalen baseline cumulative hazard estimates for mortality outcomes. Imputation was performed exclusively on validation datasets using the "mice: Multivariate Imputation by Chained Equations" R package. This approach generated five complete datasets per validation cohort, maintaining statistical rigor while addressing missing data challenges.

Following predictor extraction, benchmark models were implemented with appropriate variables (e.g., generating five datasets with CHA_2_DS_2_-VASc predictors) for outcome (e.g., thromboembolic events) prediction. Predictions across the five imputed datasets were pooled through averaging for downstream analysis. This comprehensive evaluation framework was replicated for secondary bleeding outcome analyses (i.e., with models HAS-BLED and ORBIT), ensuring consistent methodological approaches across all clinical endpoints and providing robust comparative performance assessments against our TRisk model.

#### Implementation of AI models

For the TRisk class of AI models, the implementation was carried out in Python programming language using the graphical processing unit (GPU) compatible PyTorch coding framework. One NVIDIA A100 Tensor Core 80 gigabyte GPU was used for each of the model training, validation, and explainability pipelines. For optimiser, the Adam optimiser was used as conventional with the BEHRT class of models; learning rates were optimised during the course of model training using the exponential decay method.^1^

We used the All of Us (AoU) research platform for the transfer learning and validation of our AI models. We used the 8 V100 GPUs provided for use for fine-tuning and for the explainability studies.

#### Supplementary Methods: Economic impact analyses

A decision tree was constructed to compare the TRisk strategy with standard care (**Figure S13**). In the base case, the model population comprised atrial fibrillation patients initiating DOACs, with initiation rates derived from the Clinical Practice Research Datalink (CPRD) for the United Kingdom, the All of Us Research Program for the United States, and the EORP AF Long-Term General Registry for European countries. The model incorporated the probability of DOAC use, the risk of major bleeding, and 1-year mortality. Each terminal node, at the end of a clinical pathway, was assigned a cost and quality-adjusted life-year (QALY) outcome. All parameter values and data sources are summarised in **Tables S6 and S7**.

The model estimates the impact of safely reducing DOAC use under the TRisk strategy compared with standard care. Cost and outcome estimates are informed by deselection rates derived from robust real-world datasets (CPRD and All of Us) found in **S5 and S10**.

Analyses were conducted from the healthcare payer perspective over a 1-year time horizon. The model population reflected patients with atrial fibrillation initiating DOACs, informed by national age- and sex-specific prevalence estimates from the Global Burden of Disease study and initiation rates from CPRD, All of US, and the EURObservational Research Programme (EORP) AF Long-Term General Registry.

Country-specific inputs, including DOAC use, unit drug prices, bleeding-related costs, and population characteristics, were incorporated to reflect local settings. Comparable stratified data on DOAC use by age and sex were unavailable for all countries; we therefore assumed that the distribution of DOAC agents applied uniformly across age and sex groups within each country. Age- and sex-specific probabilities of 1-year mortality for UK and European countries were estimated using logistic regression models from the EORP study. Because no meaningful differences in mortality risk were observed between the UK and European regions, the same regression coefficients were applied across all countries. In the absence of comparable US-based estimates for 1-year mortality, these coefficients were also used for the United States.

Costs included drug acquisition and the management of major bleeding events, obtained from national drug price lists and published cost studies. All costs were inflated to 2024 values and converted into local currencies where necessary. QALYs were estimated using published utility values for atrial fibrillation and disutility weights for major bleeding.

Model outputs consisted of total costs, QALYs, and incremental differences between TRisk and standard care. Uncertainty in model inputs was propagated into model outputs using probabilistic sensitivity analysis. Results are presented as mean estimates with 95% credible intervals. A sensitivity analysis extending the model population to all DOAC users (both new initiators and existing users) was also conducted to estimate the upper bound of TRisk’s potential impact **(Table S10).** The model structure was identical across countries, with variation arising only from country-specific input values.

Notably, if fewer patients are ultimately selected for DOACs due to bleeding risk or other contraindications – regardless of whether CHA₂DS₂-VASc or TRisk is used – TRisk’s relative impact would increase beyond the conservative deselection rates observed (8% in the UK, 7% in the US). These estimates therefore represent a lower bound, with absolute savings remaining consistent across different clinical scenarios. The sensitivity analysis (**Table S10),** which includes all DOAC users (i.e., both new initiators and existing users), represents an upper bound on the potential impact of the TRisk strategy.

#### Supplementary Methods: Clarification on explainability analyses

In this work, we used integrated gradients to understand predictive process of the TRisk model.^2^ In gist, the integrated gradient method is a method that helps explain how AI models make decisions by tracking the importance of each input feature along a straight path from a baseline to the actual input. Starting from a neutral reference point – a zero-value vector for input to our TRisk model – it measures how much each feature contributes to the model’s final prediction by gradually transforming this baseline into the actual input. By averaging these contributions along the path, the integrated gradients method provides a clear measure of which parts of the input were most influential in the model’s decision, making complex AI models like TRisk more interpretable to humans.

In our work, we adapted the integrated gradients method to work within the SODEN framework, calculating how each patient encounter influences the model’s output. For repeated encounters (e.g., multiple diagnoses of the same condition or multiple prescriptions of same medication), we used the highest contribution value across all instances. We focused our analysis on encounters that appeared in at least 1% of patients to ensure statistical reliability. In age-based analyses, for each encounter, we grouped patients into subgroups based on age of first recording of the particular encounter. Similarly, for the analyses of time between recording of encounter and baseline, we grouped patients into subgroups based on first (as opposed to any other subsequent recording) recording of encounter and baseline.

We replicated these analyses on the cohort in All of Us dataset (see below for details on dataset).

#### Supplementary Methods: Clarification on All of Us validation study

In additional external validation analysis on the All of Us (AoU) dataset, we first identified cohort of AF patients using established ICD-10 codes for AF identification (I48 family of ICD-10 codes). We identified patients with AF at least 18 years old in the years between Jan 1 2010 and Jan 1 2023. Similar to cohort selection in CPRD, the index date (baseline) was randomly selected from the eligible patient time period for each individual that went up to the earliest of the following: last clinical visit (i.e., this is date after which patient is lost-to-follow-up), death, Jan 1 2023, and the event of interest (i.e., bleeding and thromboembolic events). Mortality data were provided for the controlled tier of data access in AoU.

On a random selection of 60% of the cohort, we conducted external validation of the TRisk model, henceforth referred to as the AoU external validation dataset. We used the remaining 40% of the identified cohort for transfer learning and fine-tuning experiments, henceforth referred to as the AoU fine-tuning dataset. Within the 40% cut of the dataset, for experiments involving TRisk model fitting/training, 10% of the dataset was randomly selected to conduct end-of-epoch testing. This testing at the end of each epoch (i.e., one full iteration of training in which the model has trained on all patient selected for training) was used to ensure the model has not overfit on training data (i.e., ultimately aiming to avoid data “memorisation”). All models including benchmark models were evaluated on the external validation dataset.

#### Supplementary Methods: Clarification on modelling on All of Us dataset

We evaluated three distinct TRisk model variants on the All of Us (AoU)^3^ validation dataset: TRisk trained exclusively on CPRD data, TRisk trained from scratch on the fine-tuning dataset, and TRisk initially trained on CPRD then transferred and fine-tuned on the fine-tuning dataset.

Since TRisk requires specific vocabulary encoding (ICD-10 for diagnoses, ingredients for medications, SNOMED for procedures), comprehensive mapping was essential for AoU dataset integration. Raw AoU records underwent systematic conversion to match CPRD vocabulary standards, enabling successful model transfer between UK and US healthcare systems. We utilized established OHDSI Athena mapping files to convert diverse coding formats into our standardized vocabularies.

The AoU dataset contained numerous codes absent from the UK dataset, necessitating innovative vocabulary expansion strategies. For the transferred and fine-tuned TRisk model, we implemented "ontology-driven vocabulary and embedding mapping" to address this challenge. Our approach first identified codes occurring in at least 5% of AoU records. If codes existed within our established 8,675-code TRisk dictionary, no modifications were required. For missing codes, we identified the closest ontological matches. For example, if "I53.4" was absent but "I53.5" existed, we designated "I53.5" as the closest match, then added "I53.4" to the TRisk dictionary.

Importantly, rather than leaving such new code embeddings uninitialized, we loaded embeddings from the closest existing vocabulary codes (e.g., using "I53.5" embeddings for "I53.4"). For medication ingredients and procedures, we employed "fuzzy" semantic content matching to identify closest analogues. This methodology enabled effective model transfer to US data while leveraging learned representations. In this way, we conducted internal embedding transfers within the model architecture, ensuring new codes without readily available embeddings received appropriate initialization based on semantically similar existing codes.

This comprehensive approach facilitated robust cross-system model transfer, leveraging pre-trained CPRD knowledge while adapting to AoU-specific vocabulary and coding patterns. The ontology-driven strategy preserved semantic relationships between medical concepts across different healthcare systems, enabling effective model adaptation for US data while maintaining the benefits of extensive UK training data.

For the benchmark models, we implemented CHA_2_DS_2_-VASc and HAS-BLED. We did not implement the ORBIT model as this model is not used in the US. Similar to analyses on the CPRD dataset, imputation was conducted for missing variables for the HAS-BLED predictor set.

## Supplementary Figures

**Figure S1.**
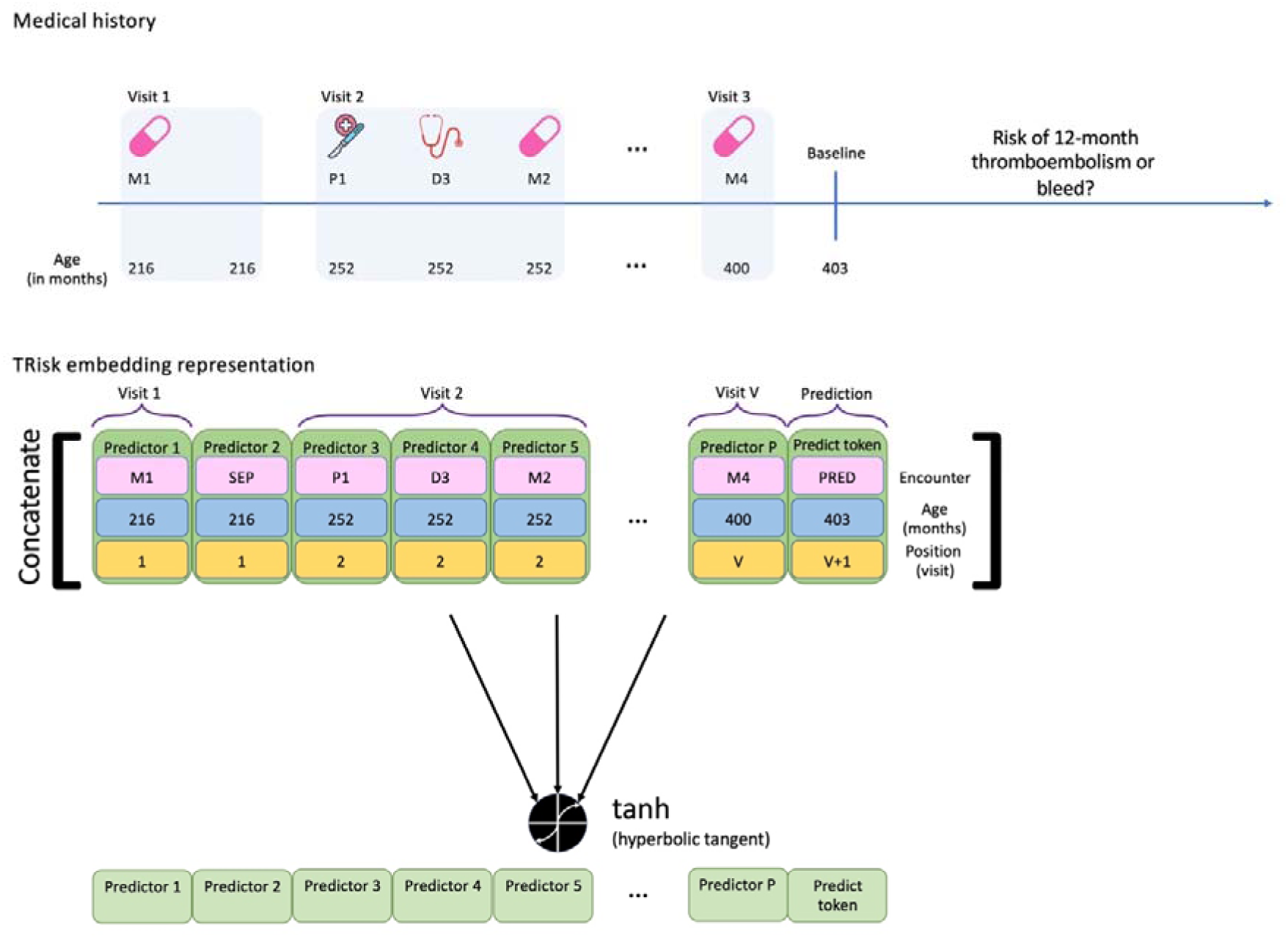
Hypothetical patient medical history and TRisk input space for this patient. For a hypothetical patient, the top part of the figure presents a sequence of medical history records represented by a group of three variables. Each record is comprised of an encounter (e.g., “M1” representing a hypothetical medication), the age at which the encounter was recorded in months (e.g., “216” months), and the visit number (e.g., visit “1”). Baseline or study entry is denoted by vertical line in the medical history timeline with vertical blue line that is when patient is 403 months old. In the bottom, we present that this medical history encounters (e.g., M1 at visit 1) in the raw electronic health record data are represented by embeddings inputted into the model. The first character of the encounter code, D, M, and P represent diagnosis, medication, and procedure records respectively represented by a code in this figure. The number following the letter illustrates a hypothetical code of that modality type (e.g., ‘D3’ might represent code “I51.2” in ICD-10 encoding). “SEP” represents a separation character given to the model to split up the data between visits. Lastly, the “PRED” token (i.e., “predict token”) is a special token that is used for prediction of the outcome (e.g., all-cause mortality outcome prediction). This special token has the age at baseline (e.g., “403” months at baseline for this hypothetical patient). As this is the visit that will happen following the final visit (i.e., visit number “V”), the visit for the predict token is appropriately enumerated as visit number “V+1”. This predict token is inputted as displayed and following further transformation by Transformer architecture layers, the corresponding output state token will be used as a condensed latent patient representation layer for input into the ordinary differential equation-based survival prediction network layers.

**Figure S2.**
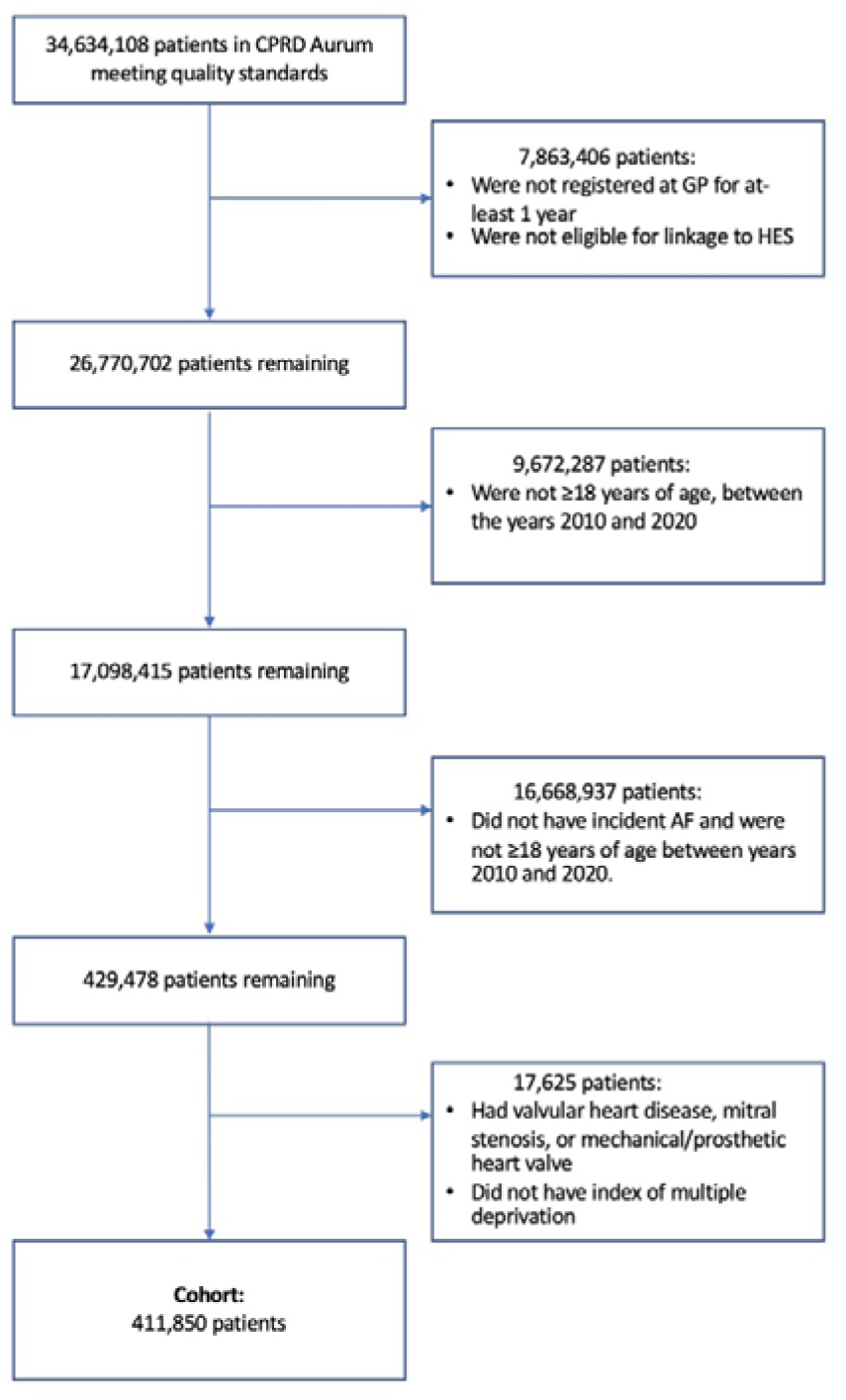
Cohort selection flowchart for Clinical Practice Research Datalink (CPRD) Aurum dataset.

**Figure S3.**
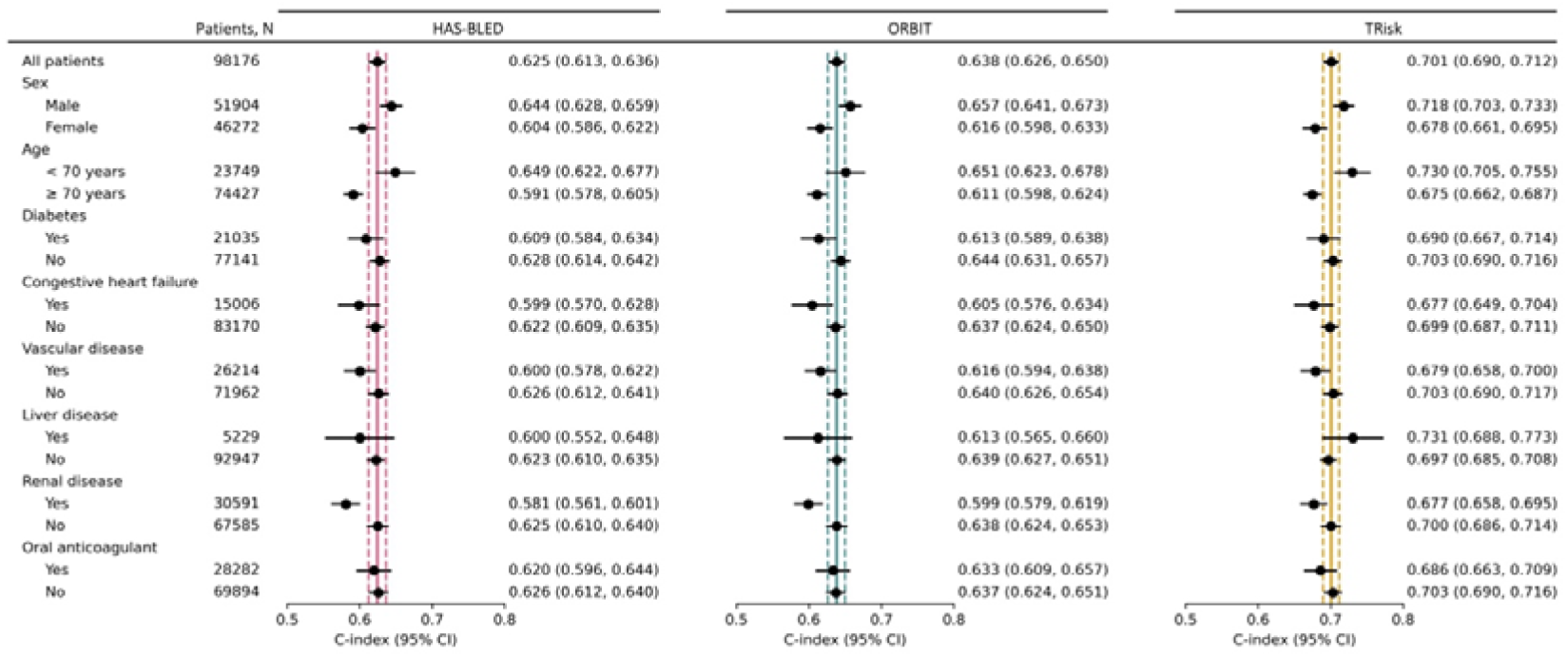
Models’ discrimination by concordance index (C-index) and associated 95% confidence intervals (CI) in overall cohort and subgroups for 12-month bleeding event risk prediction on UK validation data. Maroon, teal, and gold lines represent C-index on “all patients” in the validation cohort for HAS-BLED, ORBIT, and TRisk respectively. These solid and dotted coloured lines represent mean and 95% confidence interval boundaries of the overall cohort C-index for each model. These lines are provided to visually demonstrate deviation in subgroup discrimination performance from overall cohort performance for each model. SBP: systolic blood pressure.

**Figure S4.**
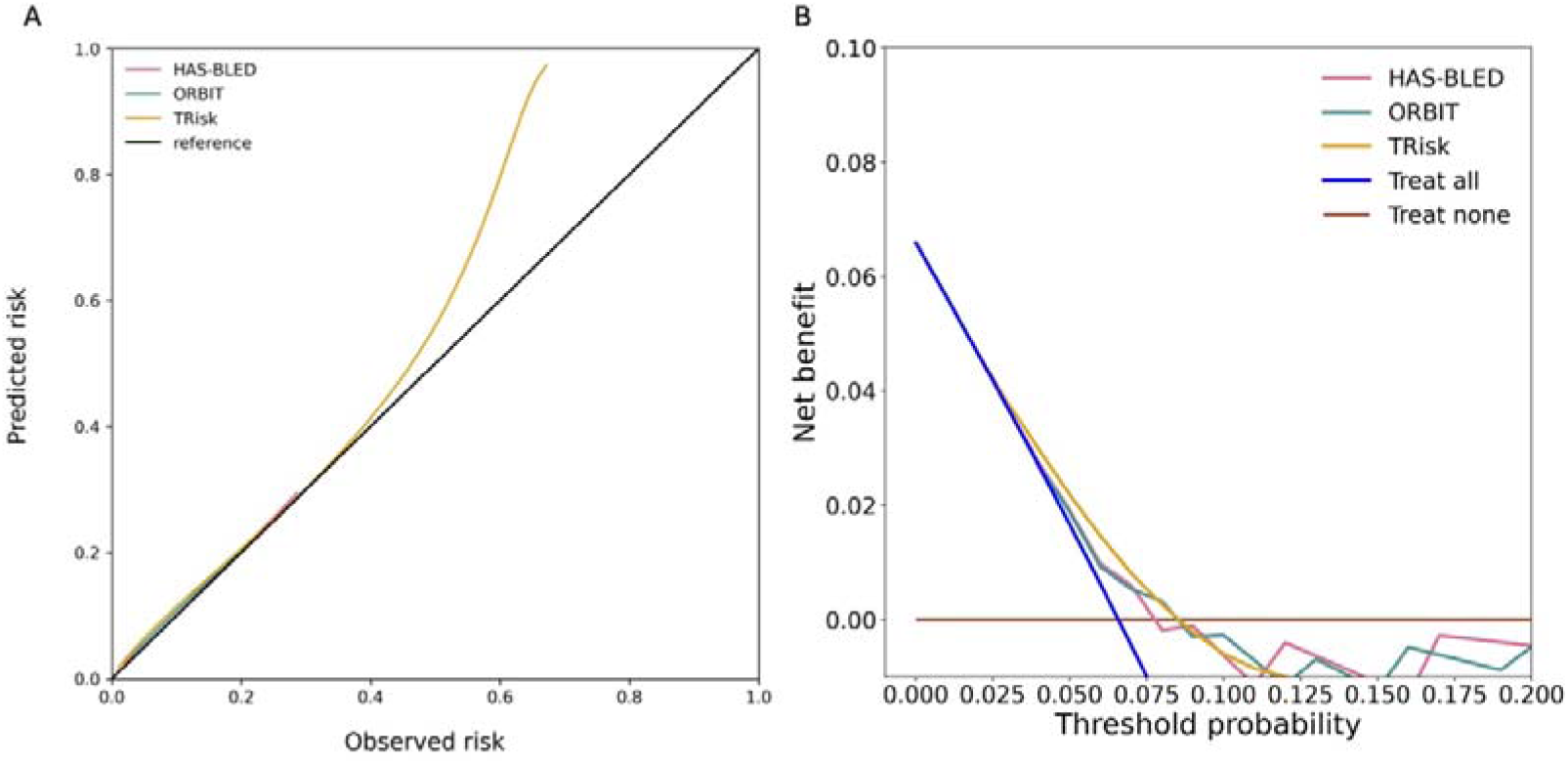
Calibration curves and decision curve analyses for 12-month bleeding event prediction across all models on UK validation data. (A) Calibration curves and (B) decision curve analyses are presented for all models. Decision curve analysis (including censored observations) has been conducted for all models. Threshold probability is shown on the x-axis and the net benefit, a function of threshold probability, is shown on the y-axis and is the difference between the proportion of true positives and false positives weighted by odds of the respective decision threshold.

**Figure S5.**
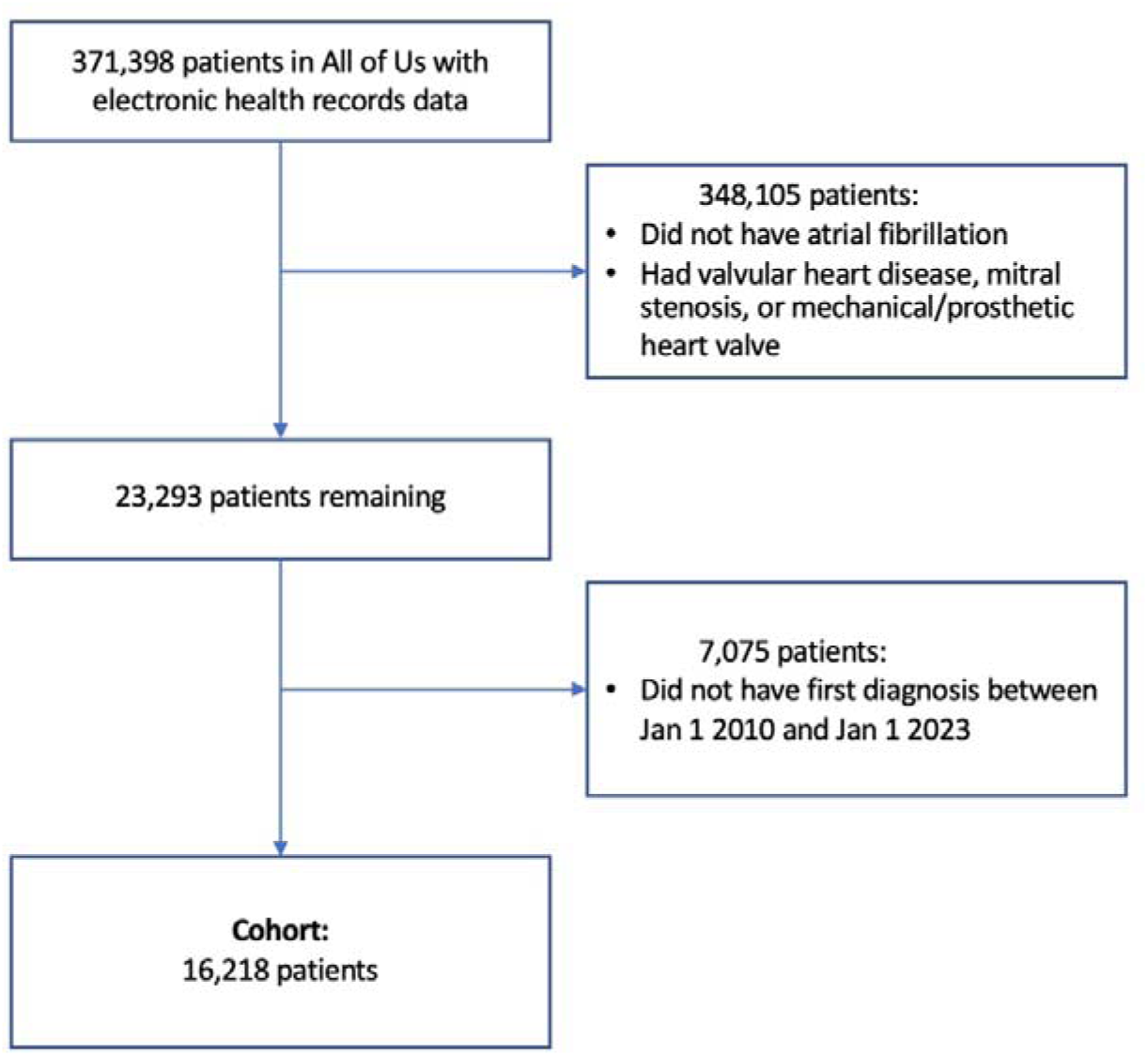
Cohort selection flowchart for All of Us (AoU) dataset.

**Figure S6.**
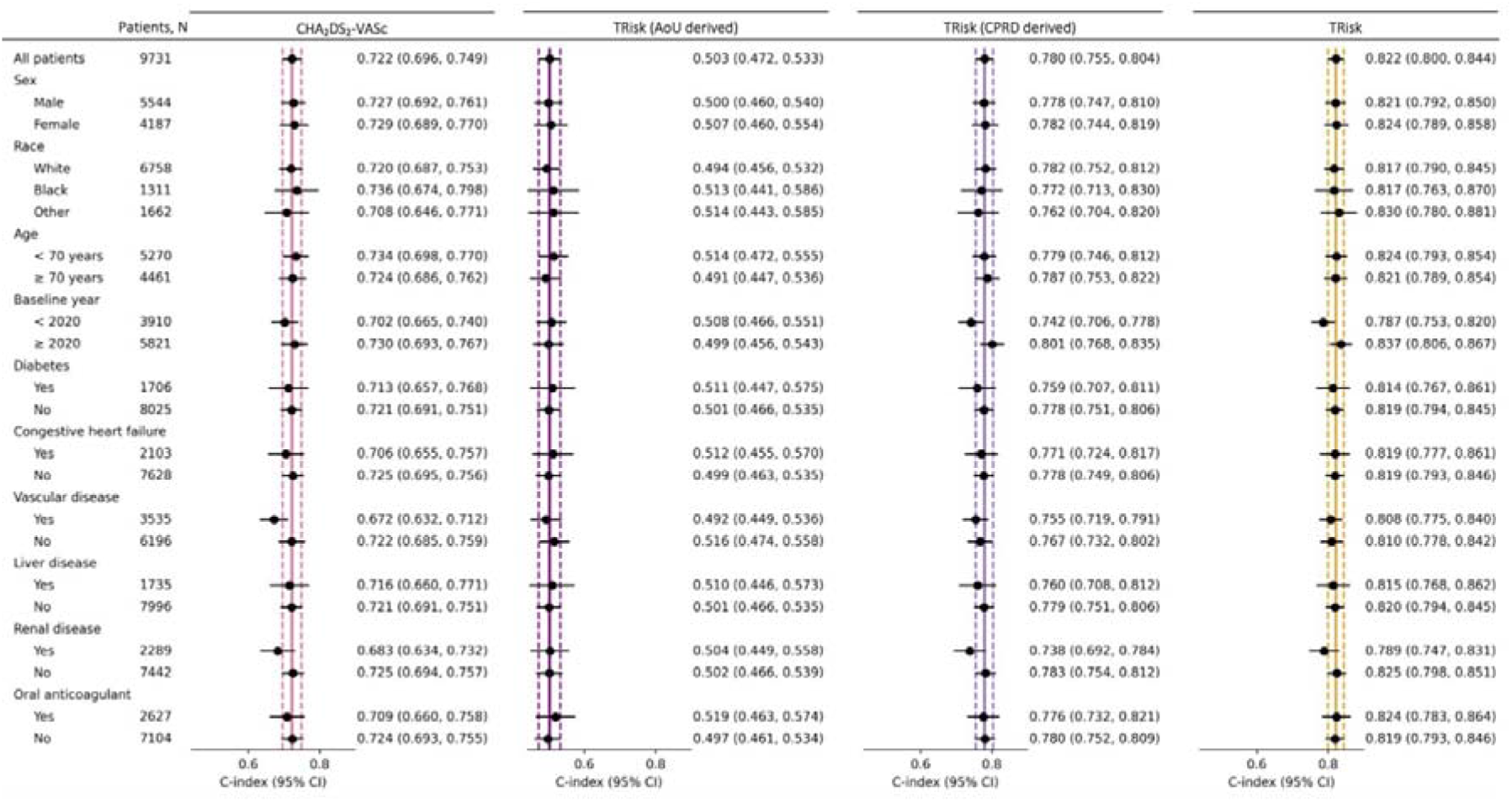
Models’ discrimination by concordance index (C-index) and associated 95% confidence intervals (CI) in overall cohort and subgroups for 12-month thromboembolic event risk prediction on US validation data. Maroon, violet, lilac, and gold lines represent C-index on “all patients” in the validation cohort for CHA_2_DS_2_-VASc, TRisk (AoU derived), TRisk (CPRD derived), and TRisk (i.e., using transfer learning) respectively on All of Us (US) validation data. These solid and dotted coloured lines represent mean and 95% confidence interval boundaries of the overall cohort C-index for each model. These lines are provided to visually demonstrate deviation in subgroup discrimination performance from overall cohort performance for each model. SBP: systolic blood pressure.

**Figure S7.**
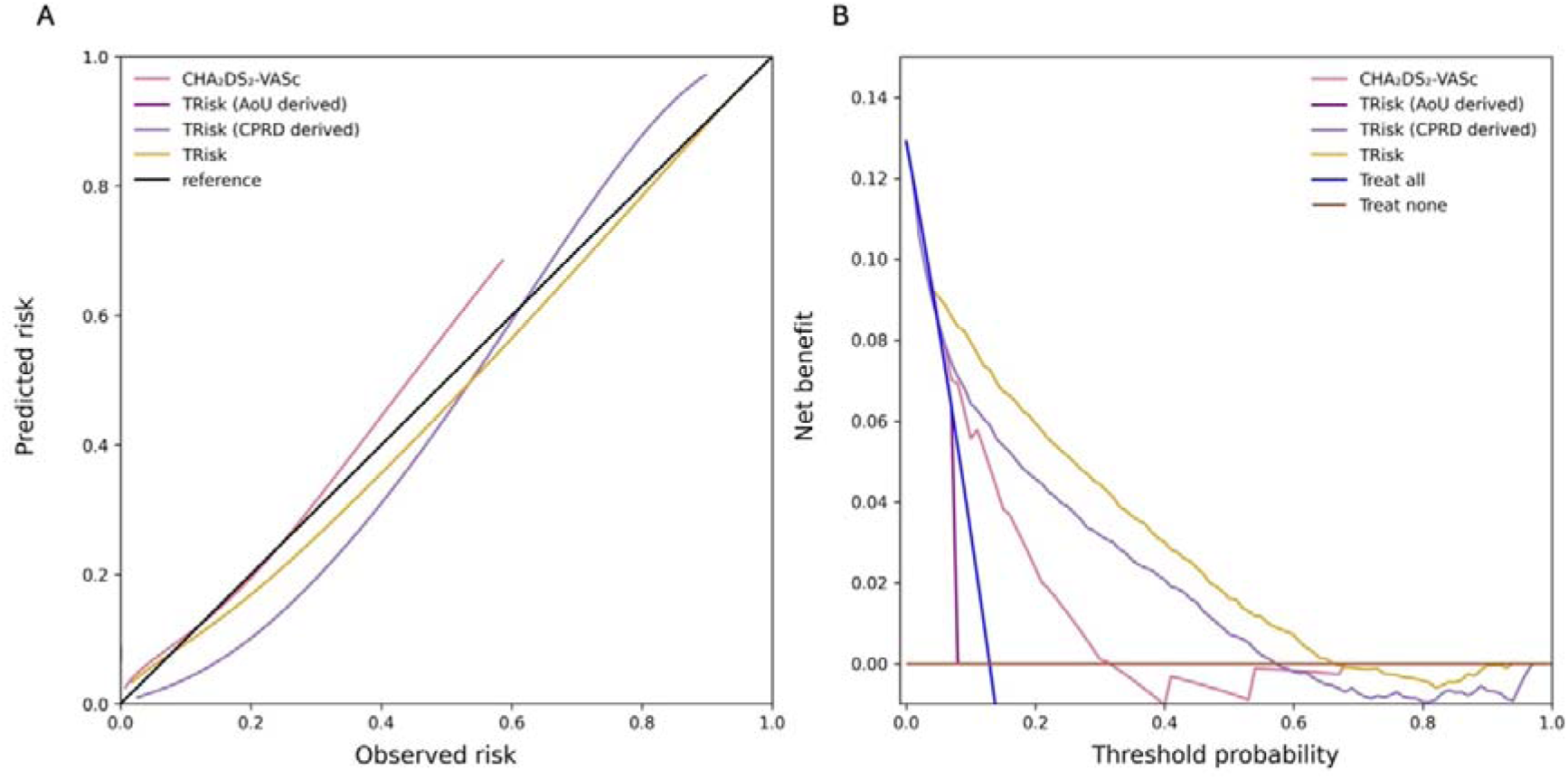
Calibration curves and decision curve analyses for 12-month thromboembolic event prediction across all models on US validation data. (A) Calibration curves and (B) decision curve analyses are presented for all models for thromboembolic event prediction on All of Us (US) validation data. Decision curve analysis (including censored observations) has been conducted for all models. Threshold probability is shown on the x-axis and the net benefit, a function of threshold probability, is shown on the y-axis and is the difference between the proportion of true positives and false positives weighted by odds of the respective decision threshold.

**Figure S8.**
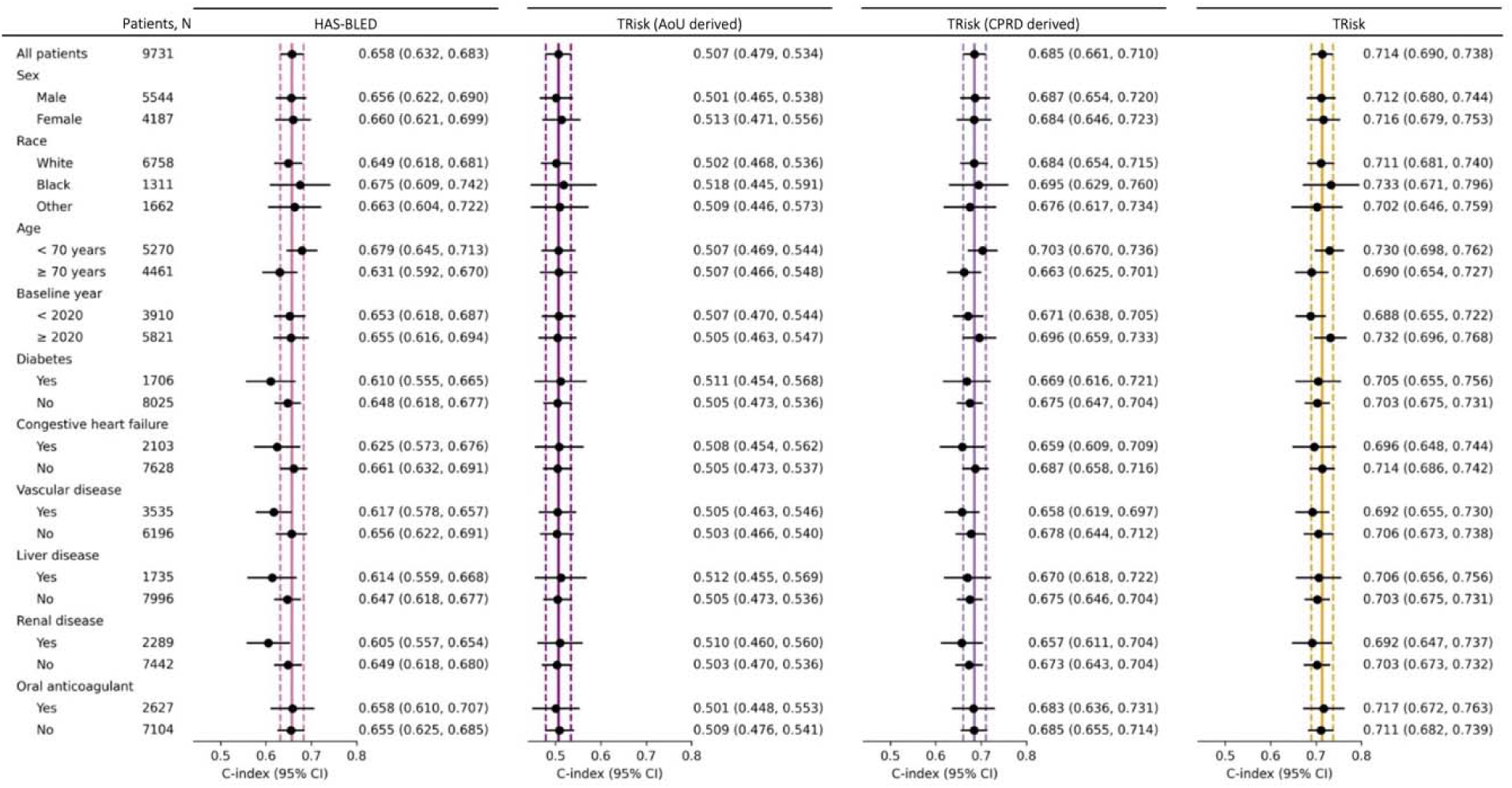
Models’ discrimination by concordance index (C-index) and associated 95% confidence intervals (CI) in overall cohort and subgroups for 12-month bleeding event risk prediction on US validation data. Maroon, violet, lilac, and gold lines represent C-index on “all patients” in the validation cohort for HAS-BLED, TRisk (AoU derived), TRisk (CPRD derived), and TRisk (i.e., using transfer learning) respectively on All of Us (US) validation data. These solid and dotted coloured lines represent mean and 95% confidence interval boundaries of the overall cohort C-index for each model. These lines are provided to visually demonstrate deviation in subgroup discrimination performance from overall cohort performance for each model. SBP: systolic blood pressure.

**Figure S9.**
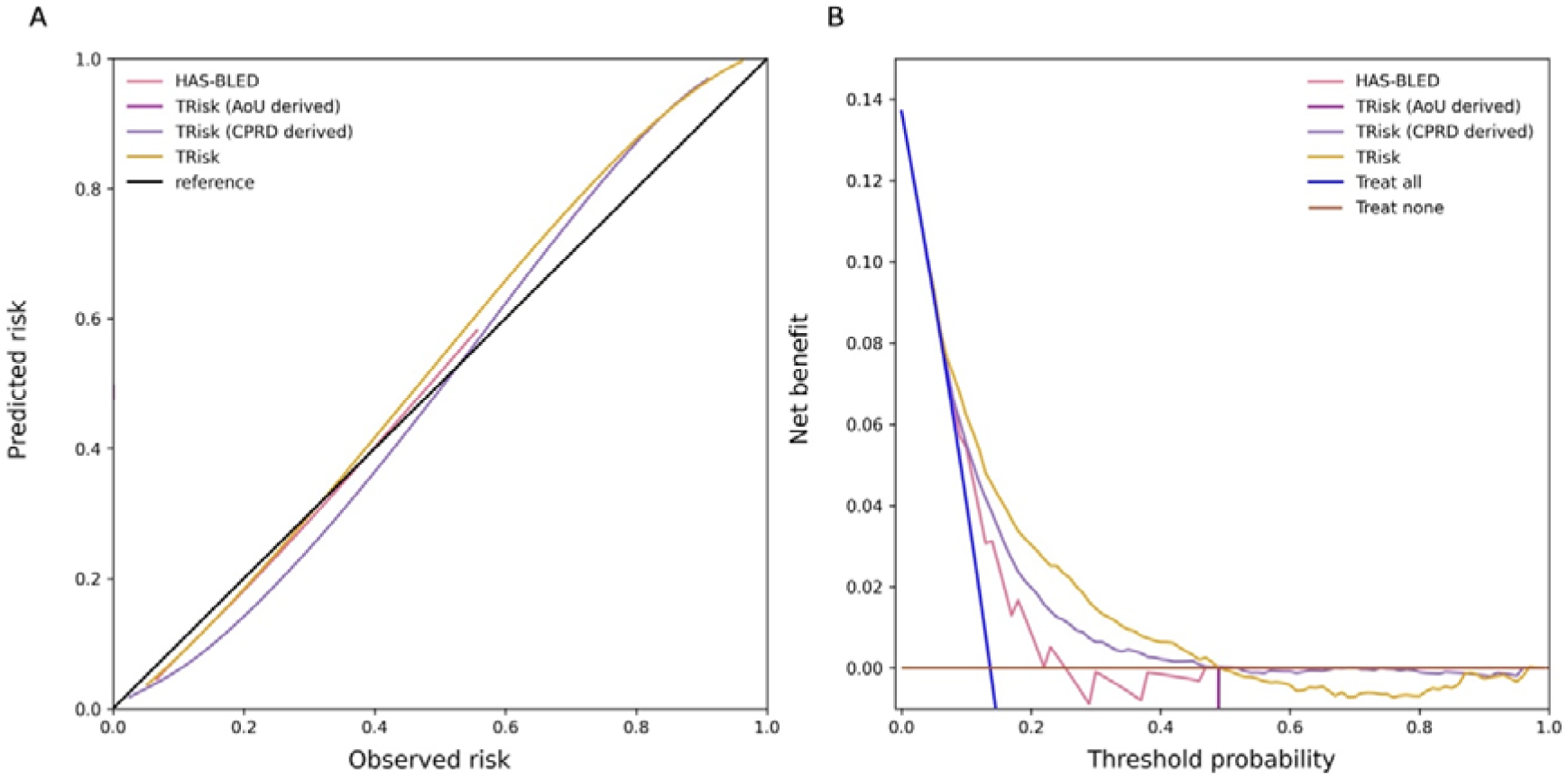
Calibration curves and decision curve analyses for 12-month bleeding event prediction across all models on US validation data. (A) Calibration curves and (B) decision curve analyses are presented for all models for bleeding event prediction on All of Us (US) validation data. Decision curve analysis (including censored observations) has been conducted for all models. Threshold probability is shown on the x-axis and the net benefit, a function of threshold probability, is shown on the y-axis and is the difference between the proportion of true positives and false positives weighted by odds of the respective decision threshold.

**Figure S10.**
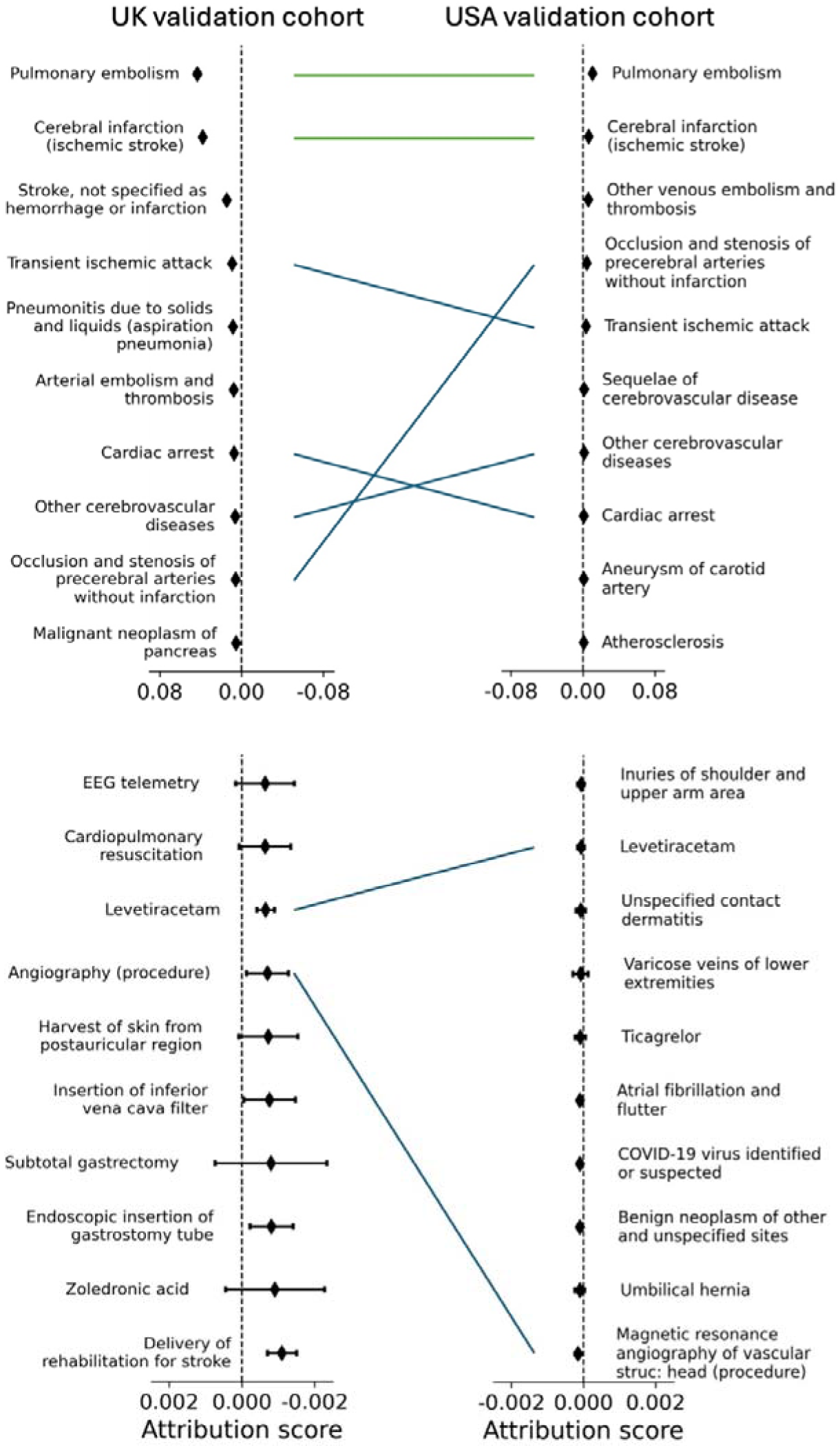
Average contribution for most positively (above) and negatively (below) contributing ten encounters for thromboembolic event risk prediction on UK and US validation cohort datasets in patients not taking oral anticoagulants. Contribution scores are captured by our TRisk model across UK (left) and US (right) validation datasets in patients not taking oral anticoagulants. Green line denotes both encounter and rank were preserved between validation analyses; blue line denotes only encounter was preserved.

**Figure S11.**
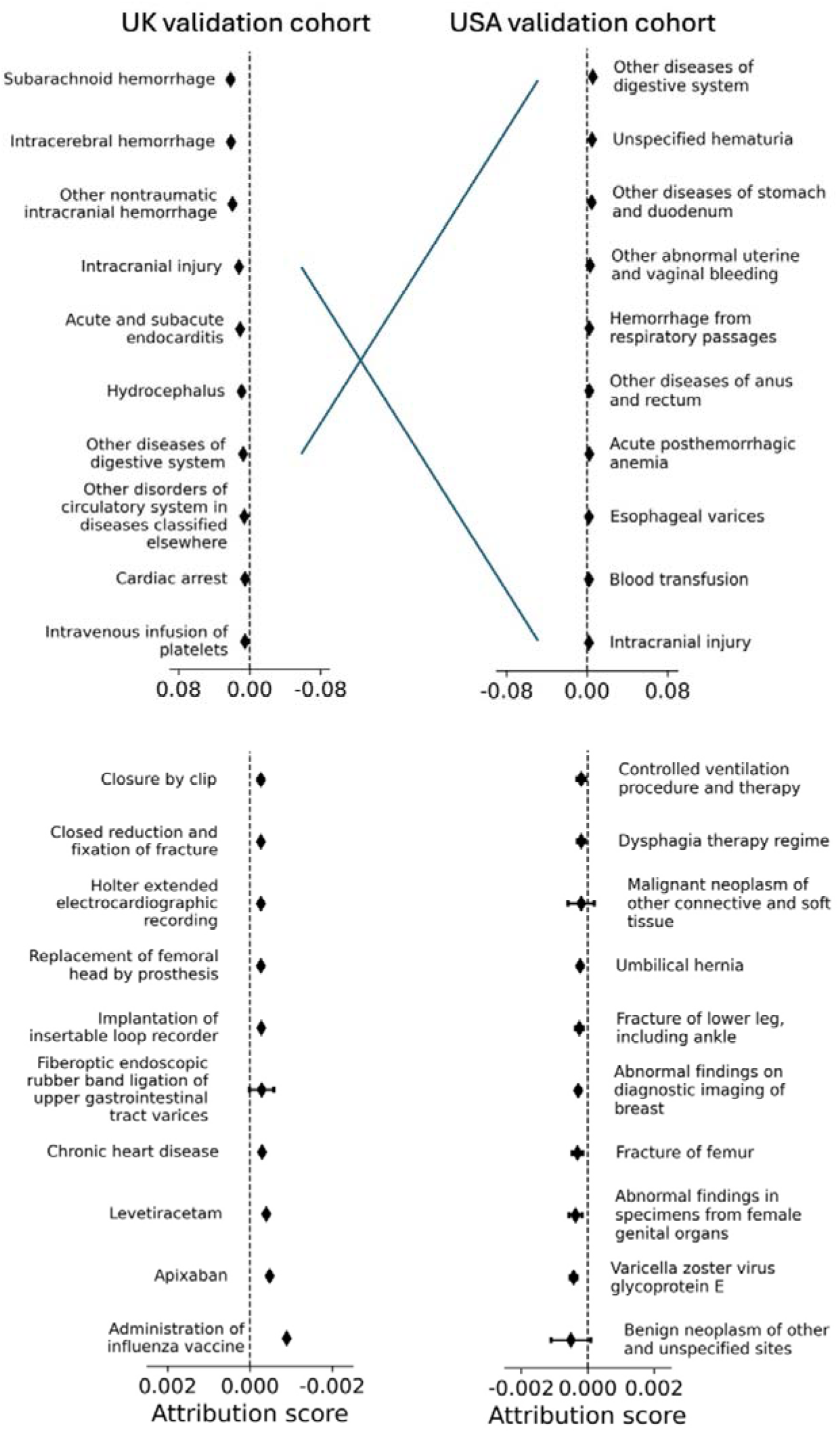
Average contribution for most positively (above) and negatively (below) contributing ten encounters for bleeding event risk prediction on UK and US validation cohort datasets. Contribution scores are captured by our TRisk model across UK (left) and US (right) validation datasets. Green line denotes both encounter and rank were preserved between validation analyses; blue line denotes only encounter was preserved.

**Figure S12.**
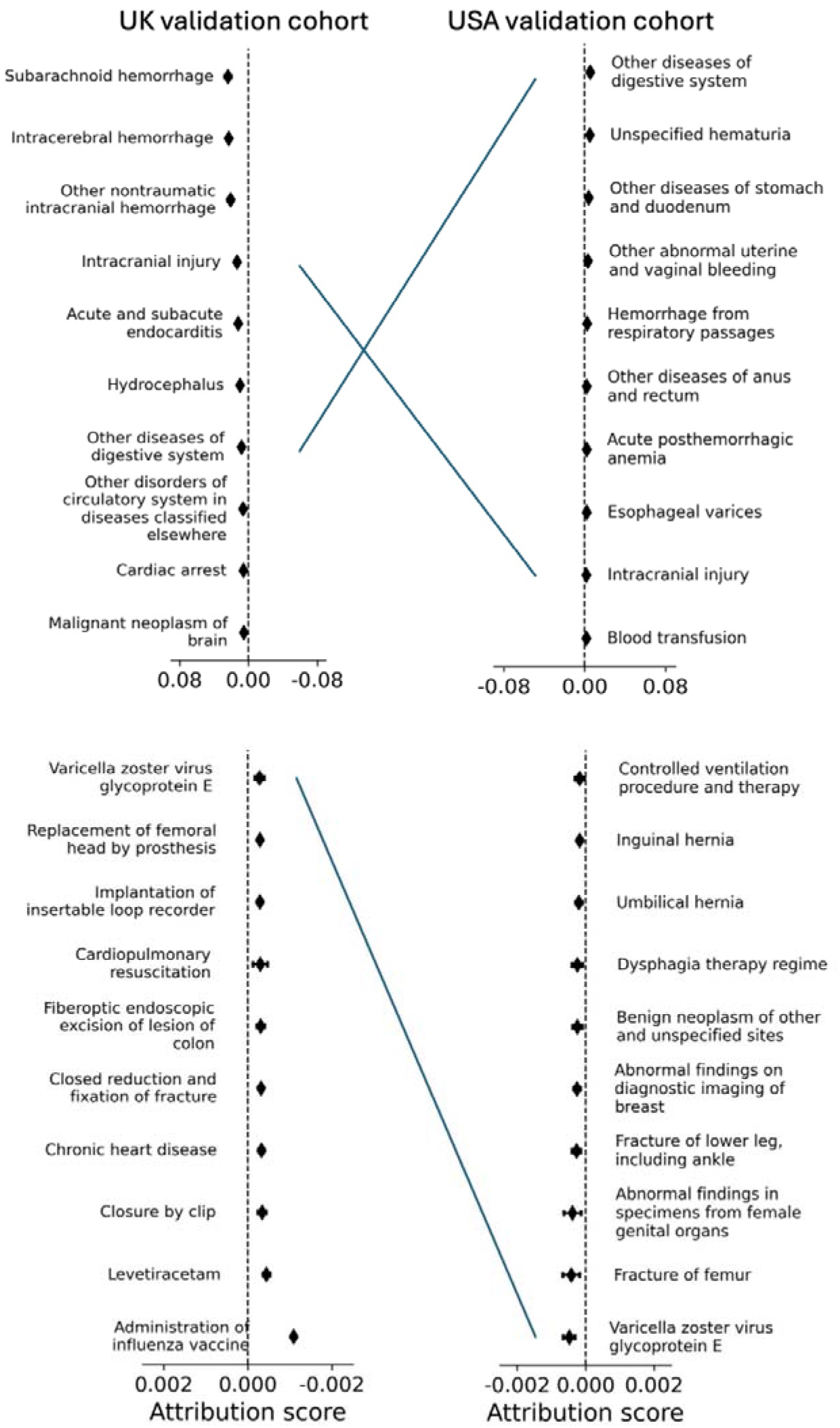
Average contribution for most positively (above) and negatively (below) contributing ten encounters for bleeding events risk prediction on UK and US validation cohort datasets in patients not taking oral anticoagulants. Contribution scores are captured by our TRisk model across UK (left) and US (right) validation datasets in patients not taking oral anticoagulants. Blue line denotes encounters were captured as important risk contributors in both cohorts.

**Figure S13.**
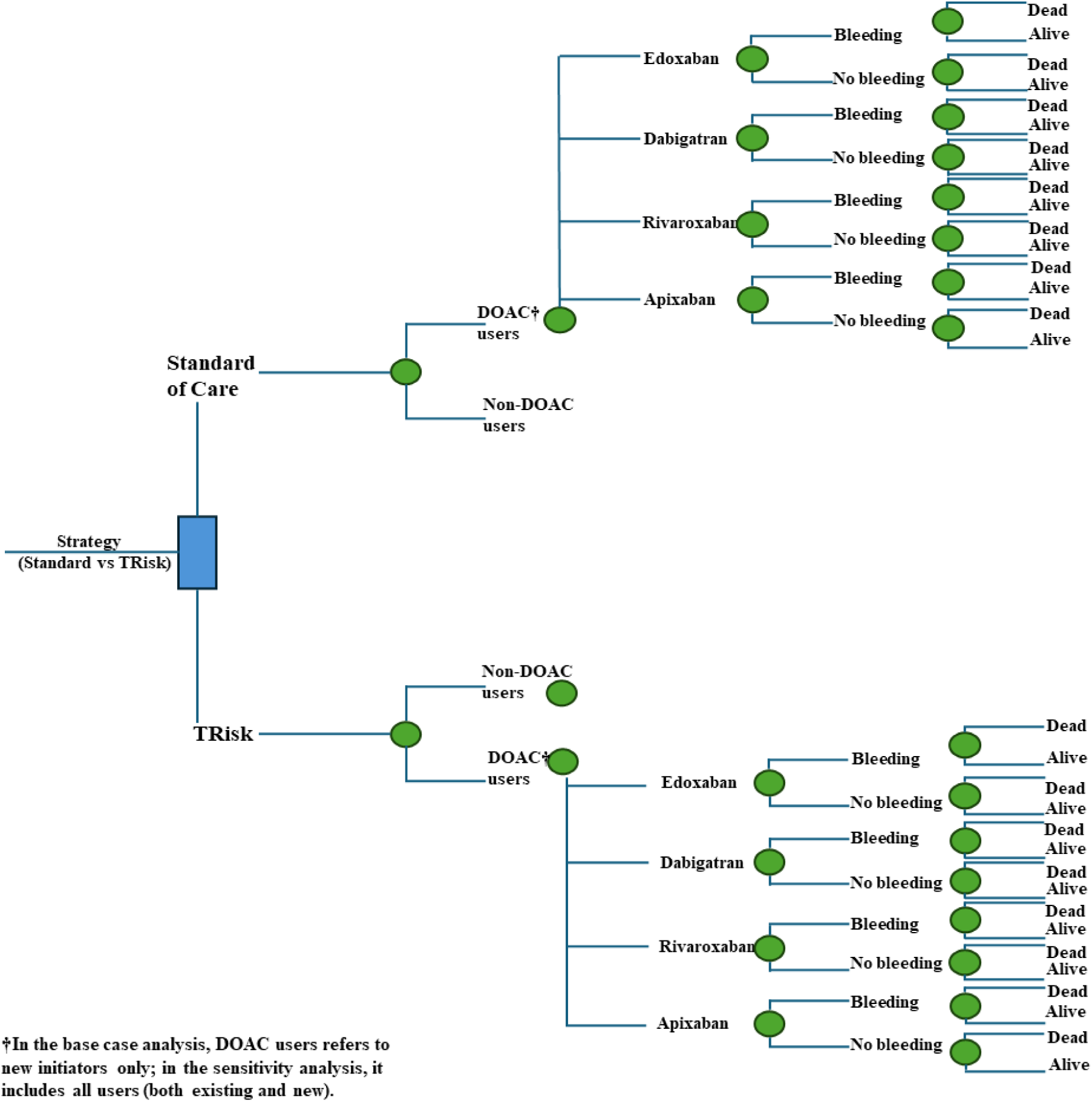
Decision tree for health economic analyses.

## Supplementary Tables

**Table S1.** Model hyperparameters and other settings for TRisk.

| Hyperparameters | Value |
| --- | --- |
| Number of layers | 7 |
| Maximum sequence length | 512 |
| Hidden size | 192 |
| Hidden dropout rate | 0.3 |
| Attention dropout rate | 0.4 |
| Number of attention heads | 6 |
| Intermediate size | 384 |
| Pooling layer size | 150 |
| Hidden activation function | Gaussian Error Linear Unit (GELU) function |
| Learning rate | 0.00003 |
| Weight decay | 0.02 |
| Warmup proportion (Adam optimiser hyperparameter) | 0.1 |
| Embedding concatenation | True |
| Embedding non-linear transformation | Hyperbolic tangent function |
| $\lambda$ weightage for explicit calibration-related modelling | 2.0 |
| Interpolation for explicit calibration-related modelling | True |
| Time bins for explicit calibration-related modelling | 48 (i.e., 1 per month) |

**Table S2.** Area under the precision-recall curve (AUPRC) metrics for all models for all 12-month risk prediction investigations on UK validation data.

|  |  | AUPRC |
| --- | --- | --- |
| Thromboembolic events | CHA<sub>2</sub>DS<sub>2</sub>-VASc | 0.123 |
|  | CHA<sub>2</sub>DS<sub>2</sub>-VA | 0.123 |
|  | TRisk | 0.238 |
|  | Model that presents chance-level performance | 0.065 |
| Bleeding events | HAS-BLED | 0.132 |
|  | ORBIT | 0.137 |
|  | TRisk | 0.158 |
|  | Model that presents chance-level performance | 0.066 |

*AUPRC: area under precision-recall curve*

**Table S3.** Population characteristics of the All of Us cohort.

| Characteristic | Derivation | Validation |
| --- | --- | --- |
| Patients, n (%) | 5,839 (36.0%) | 9,731 (60.0%) |
| Women (%) | 2,515 (43.1%) | 4,187 (43.0%) |
| Median age (years) (IQR) | 68 (61, 75) | 68 (60, 75) |
| Median SBP (mmHg) (IQR)<sup>†</sup> | 128.1 (119.7, 136.7) | 128.2 (119.6, 137.4) |
| Median creatinine (μmol/L) (IQR)<sup>†</sup> | 85.3 (72.0, 102.2) | 85.5 (72.1, 102.9) |
| Median bilirubin (μmol/L) (IQR)<sup>†</sup> | 50.1 (37.6, 69.9) | 50.7 (36.5, 70.7) |
| Drinking status (yes/no) (%)<sup>Ω</sup> | 652 (11.2%) | 1,118 (11.5%) |
| Disease history |  |  |
| Thromboembolism/stroke (%) | 1,960 (33.6%) | 3,226 (33.2%) |
| Vascular disease (%) | 2,118 (36.3%) | 3,535 (36.3%) |
| Diabetes (%) | 1,039 (17.8%) | 1,706 (17.5%) |
| Hypertension (%) | 4,604 (78.8%) | 7,687 (79.0%) |
| Congestive heart failure (%) | 1,283 (22.0%) | 2,103 (21.6%) |
| Liver disease (%) | 1,039 (17.8%) | 1,706 (17.5%) |
| CKD (%) | 1,330 (22.8%) | 2,283 (23.5%) |
| Previous bleeding (%) | 2,706 (46.3%) | 4,426 (45.5%) |
| Prescription history |  |  |
| Oral anticoagulants (%) | 1,638 (28.1%) | 2,627 (27.0%) |
| Aspirin, clopidogrel, NSAIDs (%) | 3,755 (64.3%) | 6,273 (64.5%) |

%: percent; IQR: interquartile interval; SBP: systolic blood pressure; CKD: chronic kidney disease; NSAID: non-steroidal anti-inflammatory drugs; <sup>Ω</sup> current/non-drinker status identified by ICD-10 codes; <sup>†</sup> indicates missing variables; SBP (2,484 patients; 15.3%), bilirubin (4,476; 27.6%), creatinine (2,412; 14.9%).

**Table S4.** Area under the precision-recall curve (AUPRC) metrics for all models for all 12-month risk prediction investigations on US validation data.

|  |  | AUPRC |
| --- | --- | --- |
| Thromboembolic events | CHA<sub>2</sub>DS<sub>2</sub>-VASc | 0.253 |
|  | TRisk (AoU derived) | 0.128 |
|  | TRisk (CPRD derived) | 0.423 |
|  | TRisk | 0.509 |
|  | Model that presents chance-level performance | 0.129 |
| Bleeding events | HAS-BLED | 0.215 |
|  | TRisk (AoU derived) | 0.140 |
|  | TRisk (CPRD derived) | 0.280 |
|  | TRisk | 0.342 |
|  | Model that presents chance-level performance | 0.137 |

*AUPRC: area under precision-recall curve; AoU: All of Us; CPRD: Clinical Practice Research Datalink*

**Table S5.** Impact analyses for 12-month thromboembolic event prediction on UK validation data standardised to 1,000 patients.

| Models | Threshold | Metrics |  |  |
| --- | --- | --- | --- | --- |
|  |  | TP (95% CI) | FP (95% CI) | FN (95% CI) |
| CHA<sub>2</sub>DS<sub>2</sub>-VASc | Score ≥2 for therapy recommendation (NICE 2023) | 71 (69, 73) | 814 (811, 817) | 2 (1,2) |
| CHA<sub>2</sub>DS<sub>2</sub>-VA | Score ≥3 for therapy recommendation (ESC 2024) | 70 (68, 72) | 785 (782, 788) | 3 (2,3) |
| TRisk | Risk ≥3% | 71 (69, 72) | 743 (739, 746) | 2 (1,2) |

*TP: true positive; FP: false positives; FN: false negatives; CI: confidence interval*

**Table S6.**
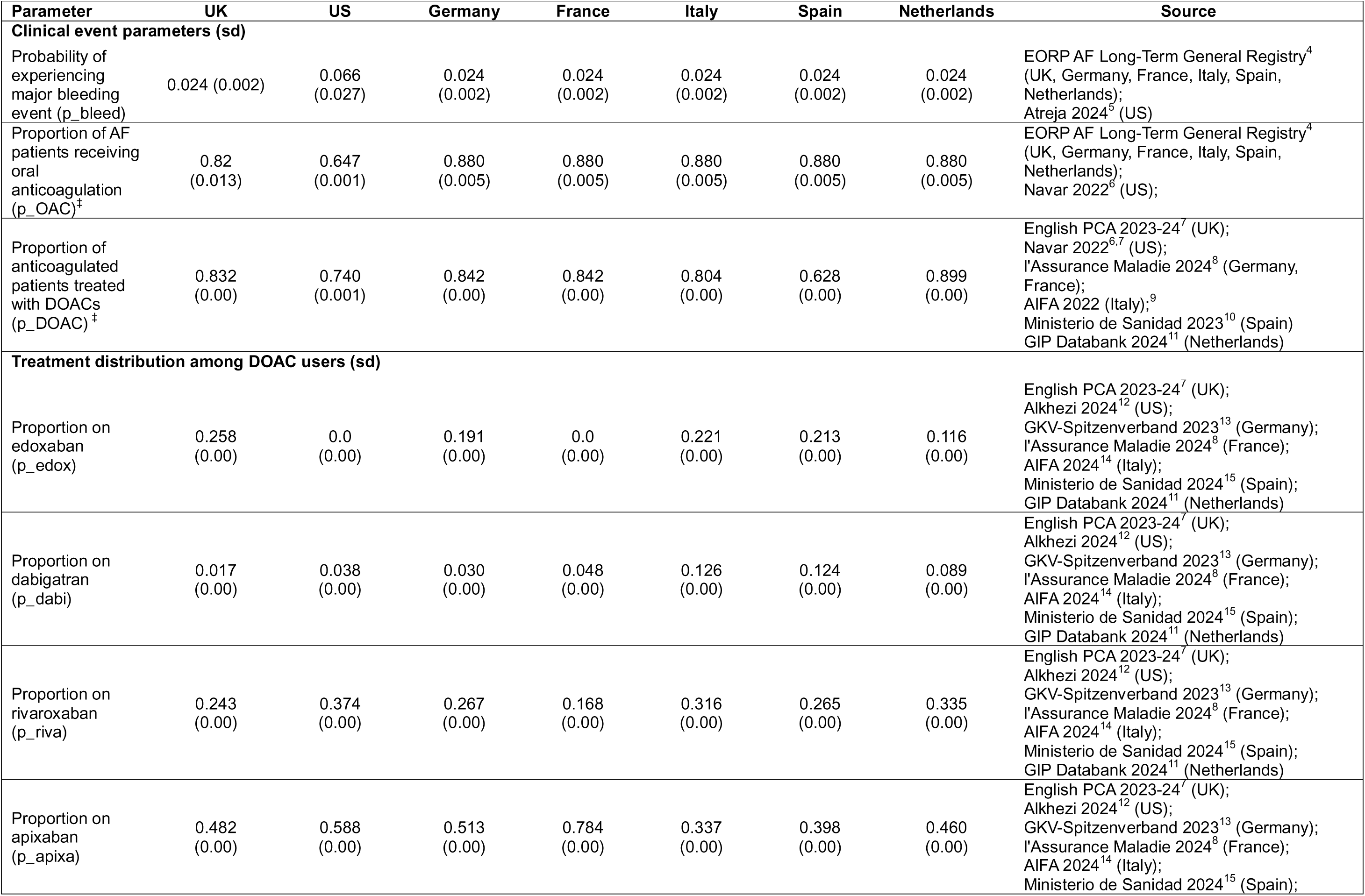
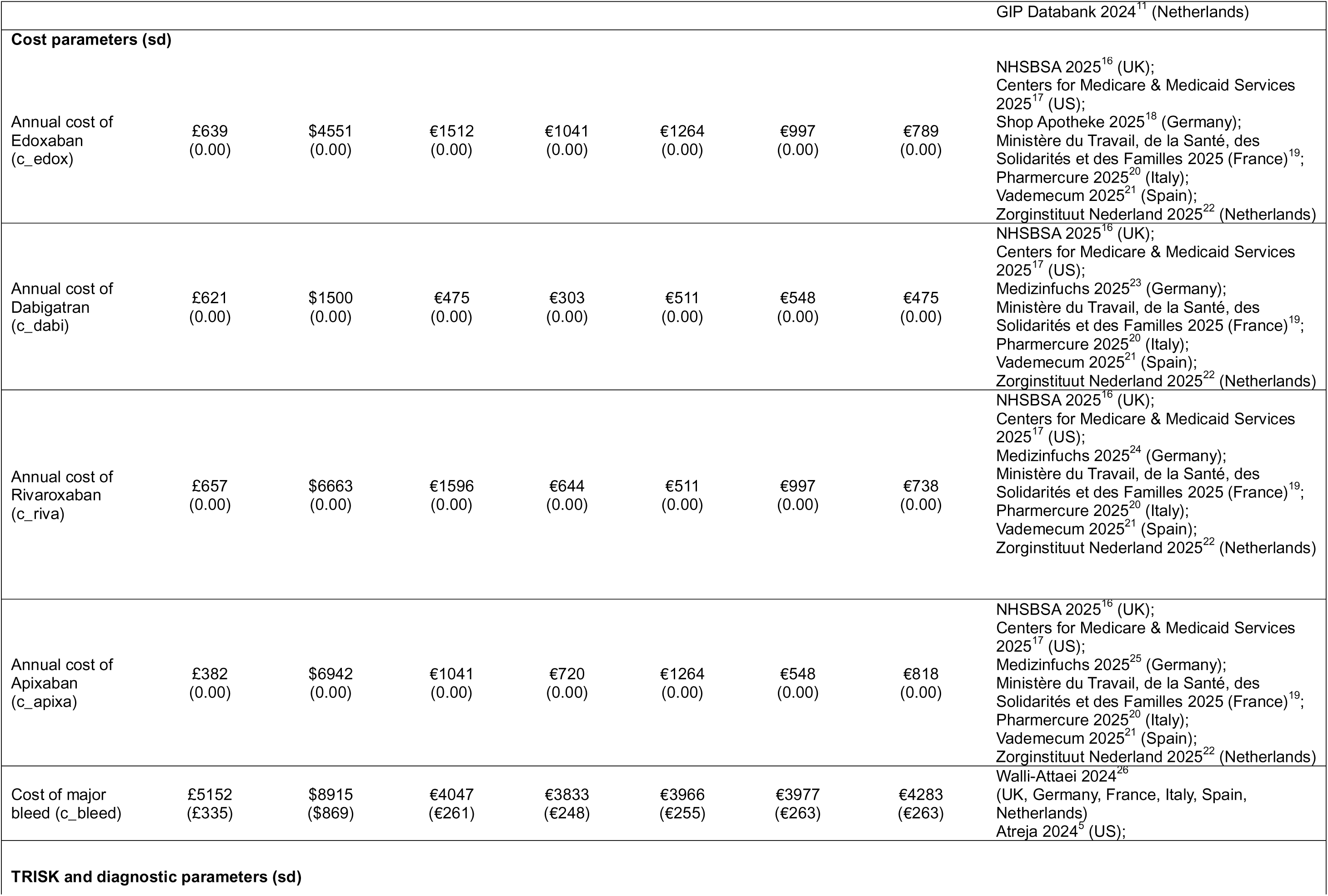
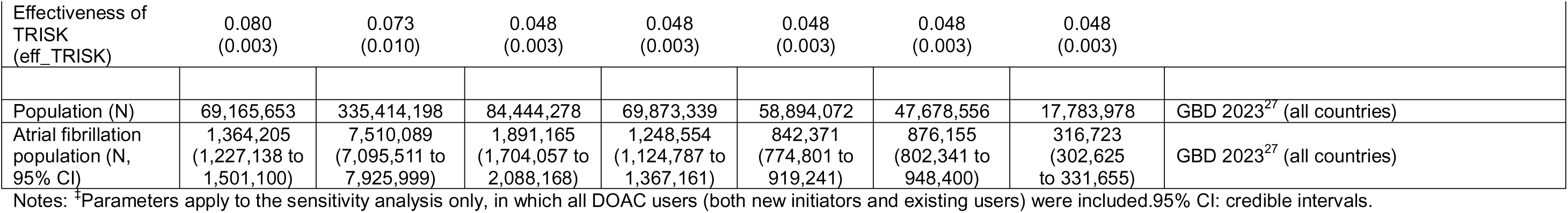
Parameters for each country and source for health economic impact evaluation.

**Table S7.** Parameters for each country and source for health economic impact analyses.

| Parameter | Value (SE) | Parameter code | Source |
| --- | --- | --- | --- |
| **Utilities** |  |  |  |
| Baseline AF utility | 0.753 (0.002) | *u_AF* | EORP AF Long-Term General Registry<sup>4</sup> |
| Utility loss from major bleed | −0.031 (0.014) | *u_bleed* | Walli-Attai 2024<sup>26</sup> |
| **DOAC use coefficients** |  |  |  |
| ***United Kingdom*** |  |  |  |
| Age | 0.002 (0.002) | *lg_doac_age* | CPRD |
| Female sex | −0.028 (0.046) | *lg_doac_female* | CPRD |
| Intercept | −2.594 (0.137) | *lg_doac_const* | CPRD |
| ***United States*** |  |  |  |
| Age | 0.025 (0.007) | *lg_doac_age* | All of Us |
| Female sex | −0.656 (0.163) | *lg_doac_female* | All of Us |
| Intercept | −3.662 (0.501) | *lg_doac_const* | All of Us |
| ***Europe*** |  |  |  |
| Age | 0.003 (0.006) | *lg_doac_age* | EORP AF Long-Term General Registry<sup>4</sup> |
| Female sex | 0.022 (0.130) | *lg_doac_female* | EORP AF Long-Term General Registry<sup>4</sup> |
| Intercept | −2.878 (0.395) | *lg_doac_const* | EORP AF Long-Term General Registry<sup>4</sup> |
| **OAC use coefficients†** |  |  |  |
| Age | 0.052 (0.003) | *lg_oac_age* | EORP AF Long-Term General Registry<sup>4</sup> |
| Female sex | −0.084 (0.060) | *lg_oac_female* | EORP AF Long-Term General Registry<sup>4</sup> |
| UK indicator | −0.181 (0.086) | *lg_oac_uk* | EORP AF Long-Term General Registry<sup>4</sup> |
| Intercept | −1.796 (0.166) | *lg_oac_const* | EORP AF Long-Term General Registry<sup>4</sup> |
| **Mortality coefficients** |  |  |  |
| Age | 0.078 (0.005) | *lg_mort_age* | EORP AF Long-Term General Registry<sup>4</sup> |
| Female sex | −0.085 (0.092) | *lg_mort_female* | EORP AF Long-Term General Registry<sup>4</sup> |
| Intercept | −8.571 (0.413) | *lg_mort_const* | EORP AF Long-Term General Registry<sup>4</sup> |

Notes: †Parameters apply to the sensitivity analysis only, in which all DOAC users (both new initiators and existing users) were included.

**Table S8.** Maximum per-patient cost at which TRisk remains cost-neutral, by country (mean and 95% credible interval) for health economic impact analyses.

| Country | Mean (95% CI) |
| --- | --- |
| United Kingdom | £50 (£45 to £54) |
| United States | \$508 (\$362 to \$650) |
| Germany | €63 (€54 to €71) |
| France | €36 (€31 to €40) |
| Italy | €47 (€41 to €53) |
| Spain | €40 (€35 to €44) |
| Netherlands | €40 (€35 to €45) |

**Table S9.** Estimated health economic impact of TRisk versus standard care across seven countries: mean values with 95% credible intervals: base case.

|  | Initiating DOACs | Number of bleeds | Cost of DOACs (million, M) | Cost of bleeds (million, M) | QALYs gained | Total costs (million, M) |
| --- | --- | --- | --- | --- | --- | --- |
| **United Kingdom** |  |  |  |  |  |  |
| SoC | 110,404 | 2,629 | £54.9 M | £13.5 M | -78 | £68.5 M |
| TRisk | 101,547 | 2,418 | £50.5 M | £12.5 M | -72 | £63.0 M |
| **Δ (95% CI)** | **-8,857**<br>**(-10,005 to -7,822)** | **-211**<br>**(-247 to -170)** | **-£4.4 M (-£5.0 M to -£3.9 M)** | **-£1.1 M (-£1.3 M to -£0.8 M)** | **6 (1 to 12)** | **-£5.5 M (-£6.2 M to -£4.8 M)** |
| **United States** |  |  |  |  |  |  |
| SoC | 897,387 | 59,228 | \$5,704.3 M | \$528.0 M | -1,760 | \$6,232.3 M |
| TRisk | 831,698 | 54,892 | \$5,286.8 M | \$489.4 M | -1,632 | \$5,776.1 M |
| **Δ (95% CI)** | **-65,688 (-85,594 to -46,481)** | **-4,335 (-8,337 to -1,537)** | **-\$417.6 M (-\$545.8 M to -\$295.0 M)** | **-\$38.7 M (-\$78.0 M to -\$13.3 M)** | **129 (8 to 325)** | **-\$456.2 M (-\$602.6 M to -\$320.1 M)** |
| **Europe\*** |  |  |  |  |  |  |
| SoC | 352,477 | 8,393 | €321.9 M | €33.4 M | -249 | €355.3 M |
| TRisk | 335,574 | 7,991 | €306.4 M | €31.8 M | -237 | €338.3 M |
| **Δ (95% CI)** | **-16,902 (-18,669 to -15,201)** | **-402 (-445 to -345)** | **-€15.4 M (-€17.2 M to -€13.7 M)** | **-€1.6 M (-€1.8 M to -€1.4 M)** | **12 (6 to 17)** | **-€17.0 M (-€19.0 M to -€15.0 M)** |
| **Germany** |  |  |  |  |  |  |
| SoC | 128,779 | 3,067 | €155.6 M | €12.4 M | -91 | €168.0 M |
| TRisk | 122,603 | 2,920 | €148.2 M | €11.8 M | -87 | €160.0 M |
| **Δ (95% CI)** | **-6,175 (-7,623 to -4,912)** | **-147 (-184 to -109)** | **-€7.5 M (-€9.2 M to -€5.9 M)** | **-€0.6 M (-€0.8 M to -€0.4 M)** | **4 (0 to 8)** | **-€8.1 M (-€9.9 M to -€6.4 M)** |
| **France** |  |  |  |  |  |  |
| SoC | 85,071 | 2,026 | €55.9 M | €7.8 M | -60 | €63.7 M |
| TRisk | 80,992 | 1,929 | €53.2 M | €7.4 M | -57 | €60.6 M |
| **Δ (95% CI)** | **-4,079 (-5,053 to -3,302)** | **-97 (-121 to -75)** | **-€2.7 M (-€3.3 M to -€2.2 M)** | **-€0.4 M (-€0.5 M to -€0.3 M)** | **3 (0 to 6)** | **-€3.1 M (-€3.8 M to -€2.5 M)** |
| **Italy** |  |  |  |  |  |  |
| SoC | 57,344 | 1,366 | €51.1 M | €5.4 M | -41 | €56.5 M |
| TRisk | 54,594 | 1,300 | €48.6 M | €5.2 M | -39 | €53.8 M |
| **Δ (95% CI)** | **-2,750 (-3,367 to -2,221)** | **-65 (-83 to -49)** | **-€2.4 M (-€3.0 M to -€2.0 M)** | **-€0.3 M (-€0.3 M to -€0.2 M)** | **2 (0 to 4)** | **-€2.7 M (-€3.3 M to -€2.2 M)** |
| **Spain** |  |  |  |  |  |  |
| SoC | 59,774 | 1,423 | €43.6 M | €5.7 M | -42 | €49.3 M |
| TRisk | 56,908 | 1,355 | €41.5 M | €5.4 M | -40 | €46.9 M |
| **Δ (95% CI)** | **-2,866 (-3,507 to -2,322)** | **-68 (-85 to -51)** | **-€2.1 M (-€2.6 M to -€1.7 M)** | **-€0.3 M (-€0.3 M to -€0.2 M)** | **2 (0 to 4)** | **-€2.4 M (-€2.9 M to -€1.9 M)** |
| **Netherlands** |  |  |  |  |  |  |
| SoC | 21,509 | 512 | €15.7 M | €2.2 M | -15 | €17.8 M |
| TRisk | 20,477 | 488 | €14.9 M | €2.1 M | -15 | €17.0 M |
| **Δ (95% CI)** | **-1,031 (-1,266 to -853)** | **-25 (-31 to -19)** | **-€0.8 M (-€0.9 M to -€0.6 M)** | **-€0.1 M (-€0.1 M to -€0.1 M)** | **1 (0 to 1)** | **-€0.9 M (-€1.0 M to -€0.7 M)** |

SoC: standard of care; TRisk: TRisk-guided recommendation; DOAC: direct oral anticoagulants; QALYs: quality-adjusted life years. \*Europe represents the aggregate of Germany, France, Italy, Spain, and the Netherlands.

**Table S10.** Estimated health economic impact of TRisk versus standard care across seven countries: mean values with 95% credible intervals: sensitivity analysis.

|  | Taking DOACs<br>(thousands, K) | Number of<br>bleeds<br>(thousands, K) | Cost of DOACs<br>(million, M) | Cost of bleeds<br>(million, M) | QALYs gained | Total costs<br>(million, M) |
| --- | --- | --- | --- | --- | --- | --- |
| **United Kingdom** |  |  |  |  |  |  |
| SoC | 977.2 K | 23.3 K | £485.4 M | £119.9 M | −690.3 | £605.3 M |
| TRisk | 898.8 K | 21.4 K | £446.5 M | £110.3 M | −635.0 | £556.8 M |
| **Δ (95% CI)** | **−78.4 K (−87.9 K to −69.1 K)** | **−1.9 K (−2.2 K to −1.5 K)** | **−£38.9 M (−£43.6 M to −£34.2 M)** | **−£9.6 M (−£11.9 M to −£7.6 M)** | **55.4 (3.2 to 105.9)** | **−£48.6 M (−£54.6 M to −£42.5 M)** |
| **United States** |  |  |  |  |  |  |
| SoC | 3,597.3 K | 237.4 K | \$22,984.2 M | \$2,116.6 M | −7,093.1 | \$25,100.9 M |
| TRisk | 3,334.0 K | 220.0 K | \$21,301.8 M | \$1,961.7 M | −6,573.9 | \$23,263.5 M |
| **Δ (95% CI)** | **−263.3 K (−329.5 K to −198.9 K)** | **−17.4 K (−35.5 K to −6.3 K)** | **−\$1,682.4 M (−\$2,105.6 M to −\$1,269.1 M)** | **−\$154.9 M (−\$320.0 M to −\$53.0 M)** | **519.2 (36.2 to 1,361.4)** | **−\$1,837.4 M (−\$2,357.6 M to −\$1,376.7 M)** |
| **Europe\*** |  |  |  |  |  |  |
| SoC | 3,662.3 K | 87.2 K | €3,370.5 M | €347.7 M | −2,586.1 | €3,718.2 M |
| TRisk | 3,486.7 K | 83.0 K | €3,208.9 M | €331.0 M | −2,462.1 | €3,539.9 M |
| **Δ (95% CI)** | **−175.6 K (−190.2 K to −161.2 K)** | **−4.2 K (−4.6 K to −3.7 K)** | **−€161.6 M (−€176.1 M to −€146.9 M)** | **−€16.7 M (−€18.4 M to −€14.3 M)** | **124.0 (66.8 to 178.6)** | **−€178.3 M (−€193.7 M to −€162.0 M)** |
| **Germany** |  |  |  |  |  |  |
| SoC | 1,404.0 K | 33.4 K | €1,695.3 M | €135.3 M | −991.6 | €1,830.6 M |
| TRisk | 1,336.7 K | 31.8 K | €1,614.0 M | €128.8 M | −944.0 | €1,742.8 M |
| **Δ (95% CI)** | **−67.3 K (−78.0 K to −57.0 K)** | **−1.6 K (−1.9 K to −1.3 K)** | **−€81.3 M (−€93.9 M to −€68.7 M)** | **−€6.5 M (−€7.9 M to −€4.8 M)** | **47.5 (4.3 to 89.6)** | **−€87.8 M (−€101.2 M to −€73.7 M)** |
| **France** |  |  |  |  |  |  |
| SoC | 926.6 K | 22.1 K | €608.5 M | €84.6 M | −654.2 | €693.1 M |
| TRisk | 882.1 K | 21.0 K | €579.3 M | €80.5 M | −622.8 | €659.9 M |
| **Δ (95% CI)** | **−44.4 K (−51.9 K to −37.0 K)** | **−1.1 K (−1.3 K to −0.8 K)** | **−€29.2 M (−€34.1 M to −€24.3 M)** | **−€4.1 M (−€5.0 M to −€3.1 M)** | **31.4 (4.5 to 59.5)** | **−€33.2 M (−€38.7 M to −€27.7 M)** |
| **Italy** |  |  |  |  |  |  |
| SoC | 598.1 K | 14.2 K | €532.4 M | €56.5 M | −422.1 | €588.9 M |
| TRisk | 569.4 K | 13.6 K | €506.9 M | €53.8 M | −401.8 | €560.7 M |
| **Δ (95% CI)** | **−28.7 K (−33.4 K to −24.3 K)** | **−0.7 K (−0.8 K to −0.5 K)** | **−€25.5 M (−€29.7 M to −€21.7 M)** | **−€2.7 M (−€3.3 M to −€2.0 M)** | **20.2 (1.7 to 39.1)** | **−€28.2 M (−€32.8 M to −€24.0 M)** |
| **Spain** |  |  |  |  |  |  |
| SoC | 484.4 K | 11.5 K | €353.0 M | €45.9 M | −341.6 | €398.9 M |
| TRisk | 461.2 K | 11.0 K | €336.1 M | €43.7 M | −325.2 | €379.7 M |
| **Δ (95% CI)** | **−23.2 K (−26.9 K to −19.8 K)** | **−0.6 K (−0.7 K to −0.4 K)** | **−€16.9 M (−€19.6 M to −€14.5 M)** | **−€2.2 M (−€2.6 M to −€1.7 M)** | **16.4 (2.3 to 31.1)** | **−€19.1 M (−€22.0 M to −€16.3 M)** |
| **Netherlands** |  |  |  |  |  |  |
| SoC | 249.3 K | 5.9 K | €181.3 M | €25.4 M | −176.7 | €206.7 M |
| TRisk | 237.4 K | 5.7 K | €172.6 M | €24.2 M | −168.2 | €196.8 M |
| **Δ (95% CI)** | **−12.0 K (−13.5 K to −10.4 K)** | **−0.3 K (−0.3 K to −0.2 K)** | **−€8.7 M (−€9.8 M to −€7.6 M)** | **−€1.2 M (−€1.5 M to −€0.9 M)** | **8.5 (0.2 to 15.6)** | **−€9.9 M (−€11.2 M to −€8.6 M)** |

SoC: standard of care; TRisk: TRisk-guided recommendation; DOAC: direct oral anticoagulants; QALYs: quality-adjusted life years. \*Europe represents the aggregate of Germany, France, Italy, Spain, and the Netherlands.

**Table S11.** Impact analyses for 12-month thromboembolic event prediction on US validation data standardised to 1,000 patients.

| Models | Threshold | Metrics |  |  |
| --- | --- | --- | --- | --- |
|  |  | TP (95% CI) | FP (95% CI) | FN (95% CI) |
| **CHA<sub>2</sub>DS<sub>2</sub>-VASc** | Score $\geq 2$ for men; $\geq 3$ for women<br>for therapy recommendation (AHA<br>2023) | 176 (169, 184) | 671 (662, 680) | 11 (9, 14) |
| **TRisk** | Risk $\geq 5\%$ | 176 (169, 184) | 609 (600, 619) | 11 (9, 14) |

*TP: true positive; FP: false positives; FN: false negatives; CI: confidence interval; AHA: American Heart* *Association*

## Notes

### Competing Interest Statement

SR reports institutional grants from the Medical Research Council and Oxford University Hospital NHS Trust, consulting fees from Lucem Health, and honoraria from the journal BMJ Heart. KR reports institutional research grants from the National Institute for Health Research (NIHR304997), Medical Research Council (MR/Y030419/1), European Union (101080430), Roche (R94776/CN002), British Heart Foundation (FS/PhD/22/29321), and the Novo Nordisk Oxford Big Data Partnership. KR also reports royalties and licences from Lucem Health, speaker fees from Radcliffe Cardiology, and personal fees in his capacity as Editor-in-Chief of Heart. Furthermore, he serves on a Medtronic Advisory Board with fees paid to his institution. All other authors declare no competing interests.

### Funding Statement

This study did not receive any funding

### Author Declarations

This study was approved by the CPRD Independent Scientific Advisory Committee (protocol 20_095). CPRD data are available to researchers via licence agreement following protocol approval. However, these data are not publicly available due to licensing restrictions; further details can be found on the CPRD website. NIH All of Us data are available to registered researchers through the All of Us Research Hub following completion of required training and data use agreements.

## References

1. Conrad N, Molenberghs G, Verbeke G, Zaccardi F, Lawson C, Friday JM, Su H, Jhund PS, Sattar N, Rahimi K, Cleland JG, Khunti K, Budts W, McMurray JJV. Trends in cardiovascular disease incidence among 22 million people in the UK over 20 years: population based study. BMJ. Epub ahead of print 2024. DOI: 10.1136/bmj-2023-078523.

2. Linz D, Gawalko M, Betz K, Hendriks JM, Lip GYH, Vinter N, Guo Y, Johnsen S. Atrial fibrillation: epidemiology, screening and digital health. The Lancet Regional Health - Europe 2024; 37: 100786.

3. Chao TF, Potpara TS, Lip GYH. Atrial fibrillation: stroke prevention. The Lancet Regional Health - Europe; 37. Epub ahead of print 1 February 2024. DOI: 10.1016/j.lanepe.2023.100797.

4. Joglar JA, Chung MK, Armbruster AL, Benjamin EJ, Chyou JY, Cronin EM, Deswal A, Eckhardt LL, Goldberger ZD, Gopinathannair R, Gorenek B, Hess PL, Hlatky M, Hogan G, Ibeh C, Indik JH, Kido K, Kusumoto F, Link MS, et al. 2023 ACC/AHA/ACCP/HRS Guideline for the Diagnosis and Management of Atrial Fibrillation: A Report of the American College of Cardiology/American Heart Association Joint Committee on Clinical Practice Guidelines. Circulation 2024; 149: E1–E156.

5. NICE. Atrial fibrillation NICE guidelines: Diagnosis and management of atrial fibrillation, www.nice.org.uk/guidance/ng196 (2021).

6. Van Gelder IC, Rienstra M, Bunting K V, Casado-Arroyo R, Caso V, Crijns HJGM, De Potter TJR, Dwight J, Guasti L, Hanke T, Jaarsma T, Lettino M, Løchen M-L, Lumbers RT, Maesen B, Mølgaard I, Rosano GMC, Sanders P, Schnabel RB, et al. 2024 ESC Guidelines for the management of atrial fibrillation developed in collaboration with the European Association for Cardio-Thoracic Surgery (EACTS). Eur Heart J. Epub ahead of print 30 August 2024. DOI: 10.1093/eurheartj/ehae176.

7. Chao TF, Joung B, Takahashi Y, Lim TW, Choi EK, Chan YH, Guo Y, Sriratanasathavorn C, Oh S, Okumura K, Lip GYH. 2021 Focused Update Consensus Guidelines of the Asia Pacific Heart Rhythm Society on Stroke Prevention in Atrial Fibrillation: Executive Summary *. Thromb Haemost 2021; 122: 20–47.

8. Lip GYH, Nieuwlaat R, Pisters R, Lane DA, Crijns HJGM, Andresen D, Camm AJ, Davies W, Capucci A, Olsson B, Aliot E, Cobbe S, Le Heuzey JY, Santini M, Vardas P, Manini M, Bramley C, Laforest V, Taylor C, et al. Refining clinical risk stratification for predicting stroke and thromboembolism in atrial fibrillation using a novel risk factor-based approach: The Euro Heart Survey on atrial fibrillation. Chest 2010; 137: 263–272.

9. Borre ED, Goode A, Raitz G, Shah B, Lowenstern A, Chatterjee R, Sharan L, Allen Lapointe NM, Yapa R, Davis JK, Lallinger K, Schmidt R, Kosinski A, Al-Khatib SM, Sanders GD. Predicting Thromboembolic and Bleeding Event Risk in Patients with Non-Valvular Atrial Fibrillation: A Systematic Review. Thromb Haemost 2018; 118: 2171–2187.

10. O’Brien EC, Simon DN, Thomas LE, Hylek EM, Gersh BJ, Ansell JE, Kowey PR, Mahaffey KW, Chang P, Fonarow GC, Pencina MJ, Piccini JP, Peterson ED. The ORBIT bleeding score: a simple bedside score to assess bleeding risk in atrial fibrillation. Eur Heart J 2015; 36: ehv476.

11. Pisters R, Lane DA, Nieuwlaat R, De Vos CB, Crijns HJGM, Lip GYH, Andresen D, Camm AJ, Davies W, Capucci A, Lévy S, Olsson B, Aliot E, Breithardt G, Cobbe S, Le Heuzey JY, Santini M, Vardas P, Manini M, et al. A novel user-friendly score (HAS-BLED) to assess 1-year risk of major bleeding in patients with atrial fibrillation: The euro heart survey. Chest 2010; 138: 1093–1100.

12. Yoshimura H, Providencia R, Finan C, Schmidt AF, Lip GYH. Refining the CHA2DS2VASc risk stratification scheme: shall we drop the sex category criterion? EP Europace 2024; 26: 280.

13. Lip GYH, Tran G, Genaidy A, Marroquin P, Estes C, Landsheft J. Improving dynamic stroke risk prediction in non-Anticoagulated patients with and without atrial fibrillation: Comparing common clinical risk scores and machine learning algorithms. Eur Heart J Qual Care Clin Outcomes 2022; 8: 548–556.

14. Lip GYH, Genaidy A, Tran G, Marroquin P, Estes C, Sloop S. Improving Stroke Risk Prediction in the General Population: A Comparative Assessment of Common Clinical Rules, a New Multimorbid Index, and Machine-Learning-Based Algorithms. Thromb Haemost 2021; 122: 142–150.

15. The “All of Us” Research Program. New England Journal of Medicine 2019; 381: 668–676.

16. Wolf A, Dedman D, Campbell J, Booth H, Lunn D, Chapman J, Myles P. Data resource profile: Clinical Practice Research Datalink (CPRD) Aurum. Int J Epidemiol; 48. Epub ahead of print 2019. DOI: 10.1093/ije/dyz034.

17. Rao S, Li Y, Mamouei M, Salimi-Khorshidi G, Wamil M, Nazarzadeh M, Yau C, Collins GS, Jackson R, Vickers A, Danaei G, Rahimi K. Refined selection of individuals for preventive cardiovascular disease treatment with a Transformer-based risk model. Lancet Digit Health; (in press). Epub ahead of print 2025. DOI: 10.1101/2024.09.25.24314371.

18. Rao S, Ahmed N, Salimi-Khorshidi G, Yau C, Su H, Conrad N, Asselbergs FW, Woodward M, Cleland JG, Rahimi K. A Transformer-based survival model for prediction of all-cause mortality in heart failure patients: a multi-cohort study, https://arxiv.org/abs/2503.12317 (2024, accessed 19 March 2025).

19. Ajabnoor AM, Zghebi SS, Parisi R, Ashcroft DM, Faivre-Finn C, Mamas MA, Kontopantelis E. Performance of CHA2DS2-VASc and HAS-BLED in predicting stroke and bleeding in atrial fibrillation and cancer. European Heart Journal Open 2024; 4: 53.

20. Hippisley-Cox J, Coupland C, Brindle P. Development and validation of QRISK3 risk prediction algorithms to estimate future risk of cardiovascular disease: Prospective cohort study. BMJ; 357.

21. Li Y, Rao S, Solares JRA, Hassaine A, Ramakrishnan R, Canoy D, Zhu Y, Rahimi K, Salimi-Khorshidi G. BEHRT: Transformer for Electronic Health Records. Sci Rep 2020; 10: 7155.

22. S Buuren KG-O. mice: multivariate imputation by chained equations in R. J Statistical Software 2011; 45: 67.

23. Austin PC, Harrell FE, van Klaveren D. Graphical calibration curves and the integrated calibration index (ICI) for survival models. Stat Med; 39. Epub ahead of print 2020. DOI: 10.1002/sim.8570.

24. Vickers AJ, Van Calster B, Steyerberg EW. Net benefit approaches to the evaluation of prediction models, molecular markers, and diagnostic tests. BMJ (Online); 352. Epub ahead of print 2016. DOI: 10.1136/bmj.i6.

25. Sundararajan M, Taly A, Yan Q. Axiomatic attribution for deep networks. In: 34th International Conference on Machine Learning, ICML 2017. 2017.

26. Zhuang F, Qi Z, Duan K, Xi D, Zhu Y, Zhu H, Xiong H, He Q. A Comprehensive Survey on Transfer Learning. Proceedings of the IEEE.

27. Collins GS, Moons KGM, Dhiman P, Riley RD, Beam AL, Van Calster B, Ghassemi M, Liu X, Reitsma JB, Van Smeden M, Boulesteix AL, Camaradou JC, Celi LA, Denaxas S, Denniston AK, Glocker B, Golub RM, Harvey H, Heinze G, et al. TRIPOD+AI statement: Updated guidance for reporting clinical prediction models that use regression or machine learning methods. BMJ. Epub ahead of print 2024. DOI: 10.1136/bmj-2023-078378.

28. Serna MJ, Rivera-Caravaca JM, López-Gálvez R, Soler-Espejo E, Lip GYH, Marín F, Roldán V. Dynamic assessment of CHA2DS2-VASc and HAS-BLED scores for predicting ischemic stroke and major bleeding in atrial fibrillation patients. Revista Española de Cardiología (English Edition) 2024; 77: 835–842.

29. Laugesen IG, Mygind A, Grove EL, Bro F. Reasons for omitting anticoagulant treatment in patients with atrial fibrillation: an audit of patient records in general practice. BMC Primary Care 2025; 26: 166.

30. Li Y, Sperrin M, Ashcroft DM, Van Staa TP. Consistency of variety of machine learning and statistical models in predicting clinical risks of individual patients: Longitudinal cohort study using cardiovascular disease as exemplar. The BMJ; 371.

31. Orlowski A, Gale CP, Ashton R, Petrungaro B, Slater R, Nadarajah R, Cowan JC, Buck J, Smith W, Wu J. Clinical and budget impacts of changes in oral anticoagulation prescribing for atrial fibrillation. Heart 2021; 107: 47–53.

32. Huisman M V., Rothman KJ, Paquette M, Teutsch C, Diener HC, Dubner SJ, Halperin JL, Ma CS, Zint K, Elsaesser A, Bartels DB, Lip G, Abban D, Abdul N, Abelson M, Ackermann A, Adams F, Adams L, Adragão P, et al. The Changing Landscape for Stroke Prevention in AF: Findings From the GLORIA-AF Registry Phase 2. J Am Coll Cardiol 2017; 69: 777–785.

33. Kim J, Nighohossian J, Daifotis AG, He J, Shafrin J. Impact of Delayed Adoption of Novel Atrial Fibrillation Treatments. American Journal of Managed Care 2024; 30: 674–680.

34. Johansson M, Guyatt G, Montori V. Guidelines should consider clinicians’ time needed to treat. BMJ; 380. Epub ahead of print 3 January 2023. DOI: 10.1136/BMJ-2022-072953.

35. FIT FOR THE FUTURE 10 Year Health Plan for England.

## References

1. Li Y, Rao S, Solares JRA, Hassaine A, Ramakrishnan R, Canoy D, Zhu Y, Rahimi K, Salimi-Khorshidi G. BEHRT: Transformer for Electronic Health Records. Sci Rep 2020; 10: 7155.

2. Sundararajan M, Taly A, Yan Q. Axiomatic attribution for deep networks. In: 34th International Conference on Machine Learning, ICML 2017. 2017.

3. The “All of Us” Research Program. New England Journal of Medicine 2019; 381: 668–676.

4. of Cardiology (ESC) ES. EURObservational Research Programme (EORP) Atrial Fibrillation General Long-Term Registry. ESC, https://www.escardio.org/Research/registries.

5. Atreja N, Johannesen K, Subash R, Bektur C, Hagan M, Hines DM, Dunnett I, Stawowczyk E. US cost-effectiveness analysis of apixaban compared with warfarin, dabigatran and rivaroxaban for nonvalvular atrial fibrillation, focusing on equal value of life years and health years in total. J Comp Eff Res; 14. Epub ahead of print 1 January 2025. DOI: 10.57264/CER-2024-0163.

6. Navar AM, Kolkailah AA, Overton R, Shah NP, Rousseau JF, Flaker GC, Pignone MP, Peterson ED. Trends in Oral Anticoagulant Use Among 436 864 Patients With Atrial Fibrillation in Community Practice, 2011 to 2020. J Am Heart Assoc; 11. Epub ahead of print 15 November 2022. DOI: 10.1161/JAHA.122.026723.

7. Authority NHSBS. Prescription cost analysis - England 2023-24, https://www.nhsbsa.nhs.uk/prescribing-data/english-prescribing-data-epd.

8. l’Assurance Maladie. Open Medic: base complète sur les dépenses de médicaments - 2014 à 2024, https://www.assurance-maladie.ameli.fr/etudes-et-donnees/open-medic-base-complete-depenses-medicament (2025).

9. del Farmaco (AIFA) AI. Report sulle nuove prescrizioni dei farmaci inclusi in Nota 97 nel corso del 2022, sulla base delle informazioni raccolte nelle schede di prescrizione compilate attraverso il sistema Tessera Sanitaria, https://www.aifa.gov.it/documents/20142/1180832/report_prescrizioni_farmaci_Nota97_TS_2022.pdf (2023).

10. de Sanidad M. Datos mensuales y anuales de consumo de recetas médicas según clasificación ATC - Año 2023, https://www.sanidad.gob.es/areas/farmacia/consumoMedicamentos/ATC/2023.htm (2023).

11. Databank GIP. GIP Databank medication database, https://www.gipdatabank.nl (2024).

12. Alkhezi OS, Buckley LF, Fanikos J. Trends in Oral Anticoagulant Use and Individual Expenditures Across the United States from 2014 to 2020. Am J Cardiovasc Drugs 2024; 24: 433–444.

13. GKV-Spitzenverband. GKV-arzneimittel-schnellinformation für Deutschland according to § 84 para. 5 SGB V, https://www.gkv-gamsi.de/media/dokumente/quartalsberichte/2023/q3_29/Bundesbericht_GAmSi_202309_konsolidiert.pdf.

14. del Farmaco (AIFA) AI. Osservatorio Nazionale sull’impiego dei Medicinali. L’uso dei farmaci in Italia. Rapporto Nazionale Anno 2023. Roma: Agenzia Italiana del Farmaco, https://www.aifa.gov.it/documents/20142/2594020/AIFA_Rapporto%2520OsMed_2023.pdf (2024).

15. de Sanidad M. Prestación farmacéutica en el Sistema Nacional de Salud 2023: informe monográfico, https://www.sanidad.gob.es/estadEstudios/estadisticas/sisInfSanSNS/tablasEstadisticas/InfAnualSNS2023/Informe_PrestacionFarmaceutica_2023.pdf (2024).

16. (NHSBSA) NHSBSA. Dictionary of Medicines and Devices (dm+d), https://dmd-browser.nhsbsa.nhs.uk (2025).

17. NADAC (National Average Drug Acquisition Cost) 2024, https://data.medicaid.gov/dataset/99315a95-37ac-4eee-946a-3c523b4c481e (accessed 28 November 2025).

18. Apotheke S. Lixiana – Produkte und Preise, https://www.shop-apotheke.com/marken/lixiana/.

19. du Travail de la Santé des S et des F. *Base de données publique des Médicaments*, https://base-donnees-publique.medicaments.gouv.fr/ (2025).

20. Pharmercure. Pharmercure, https://www.pharmercure.com/ (2025).

21. Vademecum. Vademecum, https://www.vademecum.es/ (2025).

22. Nederland Z. Medicijnkosten, https://www.medicijnkosten.nl/.

23. Medizinfuchs.de. Dabigatranetexilat Zentiva 150 mg Hartkapseln, https://www.medizinfuchs.de/dabigatranetexilat-zentiva-150.html.

24. Medizinfuchs.de. Rivaroxaban AbZ 20 mg Filmtabletten, https://www.medizinfuchs.de/preisvergleich/rivaroxaban-abz-20-mg-filmtabletten-28-stk.-abz-pharma-gmbh-pzn-18827094.html.

25. Medizinfuchs.de. Eliquis 5 mg Filmtabletten – Preisvergleich, https://www.medizinfuchs.de/preisvergleich/eliquis-5mg-filmtabletten-200-stk.-vertriebsgemeinschaft-bristol-myers-squibb-pzn-1647809.html.

26. Walli-Attaei M, Little M, Luengo-Fernandez R, Gray A, Torbica A, Maggioni AP, Bairami F, Huculeci R, Aboyans V, Timmis AD, Vardas P, Leal J. Health-related quality of life and healthcare costs of symptoms and cardiovascular disease events in patients with atrial fibrillation: a longitudinal analysis of 27 countries from the EURObservational Research Programme on Atrial Fibrillation general lo…. Europace; 26. Epub ahead of print 1 June 2024. DOI: 10.1093/EUROPACE/EUAE146.

27. of Disease Collaborative Network GB. Global Burden of Disease Study 2023 (GBD 2023). Seattle, United States, https://ghdx.healthdata.org/ (2023).

